# Cerebral Organoids Containing an *AUTS2* Missense Variant Model Microcephaly

**DOI:** 10.1101/2022.02.23.22271091

**Authors:** Summer R. Fair, Wesley Schwind, Dominic Julian, Alecia Biel, Swetha Ramadesikan, Jesse Westfall, Katherine E. Miller, Meisam Naeimi Kararoudi, Scott E. Hickey, Theresa Mihalic Mosher, Kim L. McBride, Reid Neinast, James Fitch, Dean Lee, Peter White, Richard K. Wilson, Tracy A. Bedrosian, Daniel C. Koboldt, Mark E. Hester

## Abstract

Variants in the *AUTS2* gene are associated with a broad spectrum of neurological conditions characterized by intellectual disability, microcephaly, and congenital brain malformations. Here, we use a human cerebral organoid (CO) model to investigate the pathophysiology of a heterozygous *de novo* missense *AUTS2* variant identified in a patient with multiple neurological impairments including primary microcephaly and profound intellectual disability. Proband COs exhibit reduced growth, deficits in neural progenitor cell (NPC) proliferation and disrupted NPC polarity within ventricular zone-like regions compared to control COs. We used CRISPR-Cas9-mediated gene editing to correct this variant and demonstrate rescue of impaired organoid growth and NPC proliferative deficits. Single-cell RNA sequencing revealed a marked reduction of G1/S transition gene expression and alterations in WNT-β-Catenin signaling within proband NPCs, uncovering a novel role for AUTS2 in NPCs during human cortical development. Collectively, these results underscore the value of COs to uncover molecular mechanisms underlying AUTS2 syndrome.

## INTRODUCTION

The autism susceptibility candidate 2 (*AUTS2*) gene was first identified and found disrupted as a result of a balanced translocation event (t7;20)^1^ in a pair of monozygotic twins with autism spectrum disorder (ASD). Subsequent clinical reports suggest pathological *AUTS2* variants are more closely associated with intellectual disability (ID) rather than directly contributing to classical features associated with ASD. *AUTS2* is a highly conserved gene that spans 1.2Mb on chromosome 7q11.22 and is comprised of 19 coding exons with a predicted full-length protein of 1,259 amino acids (Fig. 1e). Biochemical studies have shown that AUTS2, in association with Casein Kinase 2 (CK2), form a polycomb repressive complex 1 (PRC1-AUTS2) which activates rather than represses transcription through recruitment of p300^2^. There are several reported isoforms of *AUTS2*, including full-length and various C-terminal isoforms, that are expressed throughout brain development^3,4^. Previous studies have shown deletions within the C-terminus isoform spanning exons 9-19 are associated with a severe neurocognitive phenotype^3,5^. Other studies have demonstrated several C-terminal isoforms, which are expressed at internal transcriptional start sites in exons 6 and 9, have critical roles during neuronal differentiation^3,4,6^.

**Figure 1.**
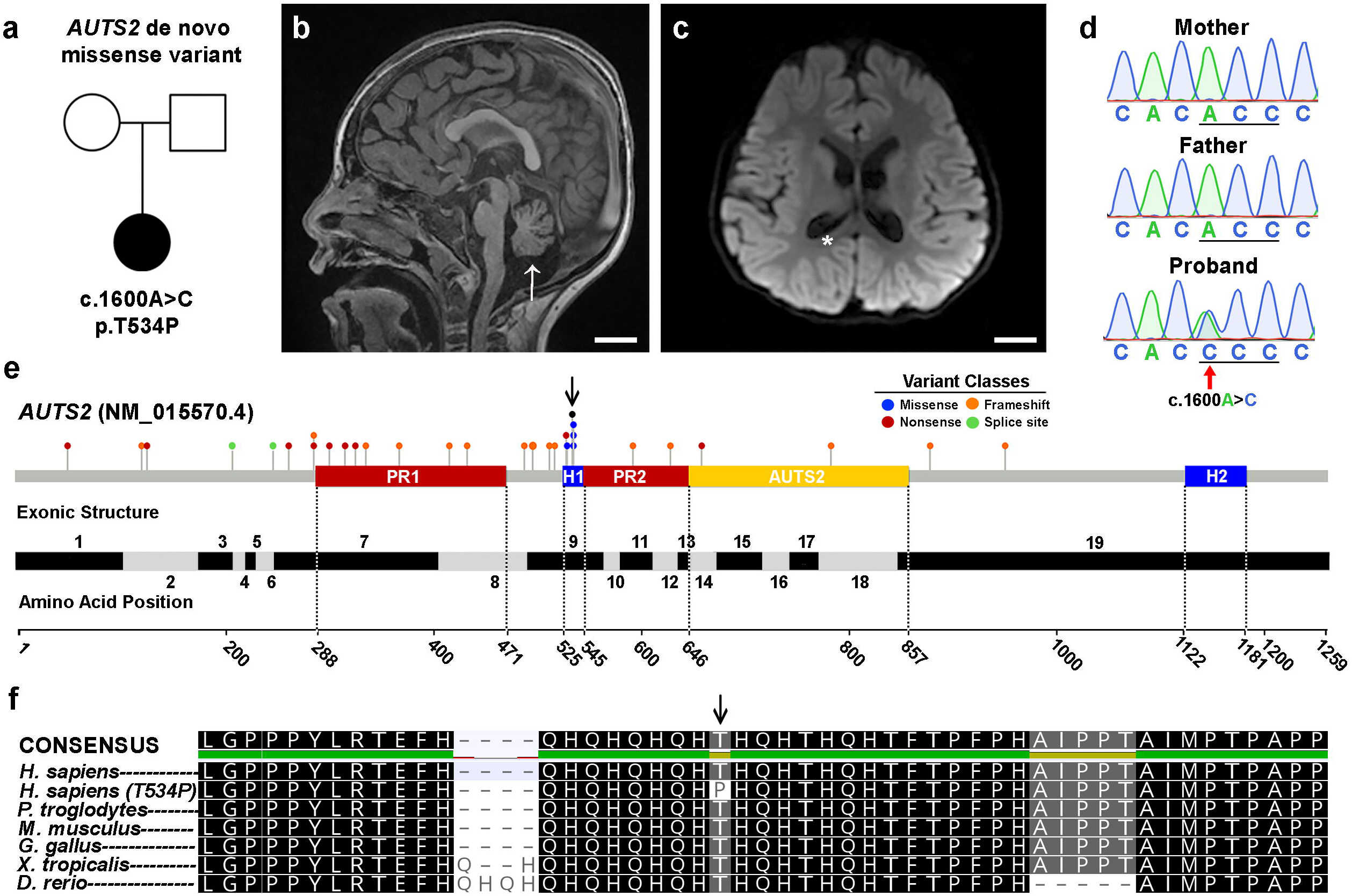
Identification of a *de novo AUTS2* variant within a conserved histidine rich domain. **(A)** Family pedigree. Circle: female; square: male; filled: affected. (**B-C)** MRI of affected individual showing reduced cortical area, ventriculomegaly (asterisk) and cerebellar atrophy (arrow). MRI images are arranged in sagittal and coronal planes, respectively. Scale bar = 2 cm. (**D)** Sanger-based DNA sequence chromatogram of the c.1600A>C *AUTS2* variant in exon 9 in the affected patient and absent in her unaffected, healthy parents. (**E)** *AUTS2* genomic organization showing disease-causing variants, key domains, and exon structure of the *AUTS2* gene. Pathogenic and likely pathogenic variants from the ClinVar database as of January 2021 are represented by circles and shaded according to variant classes. Proline-rich domains (PR1/PR2) and histidine-rich domains (H1/H2) were obtained from UniProt (entry Q8WXX7). The curated AUTS2 protein family domain from PFAM is shown in yellow. Exon locations and numbering in the bottom panel reflect the canonical full-length transcript (NM_015570.4). (**F**) AUTS2 amino acid conservation within the H1 domain and spanning position 534 across multiple species. Arrow represents the AUTS2 T534P alteration in the affected patient.

*AUTS2* variants are associated with a broad spectrum of clinical features, including low birth weight, feeding difficulties, intellectual disability, microcephaly, seizures, brain malformations and mild dysmorphic craniofacial features that are collectively known as AUTS2 syndrome^5^. *AUTS2* variants are additionally associated with a host of other neurological conditions such as addiction disorders^7,8^, epilepsy^9^, schizophrenia^10,11^, attention deficit hyperactivity disorder^12^ and dyslexia^13^. The diverse disease manifestations of *AUTS2* variants within the brain underscore the importance of elucidating its role in neurodevelopment.

Animal studies have demonstrated a putative role for AUTS2 in transcriptional activation, RNA metabolism^14^, and cytoskeletal regulation within the central nervous system (CNS) ^2,6^. *Auts2* knockdown zebrafish models exhibit overall stunted growth compared to controls, with notable reductions in the forebrain, midbrain and cerebellum^3,15^. Various rodent models of AUTS2 disruption show reductions in the cerebellum^4,6,16^ and dentate gyrus^14^. However, these rodent models do not display a microcephalic phenotype, which has hampered our understanding of molecular mechanisms that cause human microcephaly in AUTS2 syndrome. The emergence of human-based model systems of the developing brain such as cerebral organoids (COs) has refined our understanding of the mechanisms controlling human cortical development. In particular, these model systems have revealed human-specific diversity in progenitor quiescence, architecture, and subtypes that recapitulate morphological features of the developing human fetal cortex^17-20^. Several studies modeling primary microcephaly using COs arising from variants in *CDK5RAP2*^21^, *WDR62*^22^, and *NARS1*^23^ have uncovered important insights into disease mechanisms^24^. Cumulatively, these reports show cellular deficits in cell cycle progression and cilia formation in apical neural progenitor cells (NPCs) concomitant with reduced cortical expansion in COs^21-23,25,26^.

Here we describe a human CO model to investigate the pathophysiology of a heterozygous *de novo* missense *AUTS2*^*T534P*^ variant identified in a patient with several neurological impairments, including primary microcephaly and profound intellectual disability. Since the majority of all reported pathogenic/likely-pathogenic missense variants cluster near our patient’s variant, we envisioned a CO model of the *AUTS2*^*T534P*^ variant would allow us to explore disease mechanisms underlying AUTS2 syndrome. Our results demonstrate *AUTS2*^*T534P*^ COs show an impaired growth trajectory compared to controls, which recapitulates the patient’s microcephaly and thus provides a novel model to investigate causal disease mechanisms. Further investigation of proband COs using immunohistochemical techniques revealed reduced NPC proliferation and irregular apical NPC polarity in neural rosettes. To validate whether the proband’s CO phenotypes are causal to the *AUTS2*^*T534P*^ variant, we used CRISPR-Cas9 homology directed repair to restore the wild-type sequence (c.1600C>A) and demonstrate phenotypic rescue of impaired organoid growth and proliferative deficits in NPCs compared to proband COs. Single-cell RNA sequencing (scRNA-seq) of proband COs showed an underrepresentation of progenitors enriched in expression of G1/S transition genes as well as dysregulated gene expression signatures associated with WNT-β-Catenin signaling, which were rescued in gene corrected (GC) control COs. Collectively, these results demonstrate a novel role for AUTS2 during early human cortical development within NPCs and emphasize the value of COs to uncover pathogenic mechanisms underlying AUTS2 syndrome.

## RESULTS

### Identification of a de novo missense variant in the *AUTS2* gene

We report a preschool female who presented with profound intellectual disability, cerebellar hypoplasia, epilepsy, and dysmorphic features (Fig.1a). An MRI of the patient’s brain showed enlarged ventricles consistent with cerebral volume loss as well as small cerebellar hemispheres and vermis (Fig.1b-c). Extensive clinical genetic testing was performed on the patient that included karyotype and SNP microarray analysis, biochemical testing for congenital disorders of glycosylation and Smith-Lemli-Opitz syndrome, and DNA methylation testing for Prader-Willi/Angelman syndrome. All of these tests were interpreted by the clinical care team as non-diagnostic. The patient and her parents were then enrolled in an IRB-approved research study for further genomic analysis.

To investigate whether the patient’s symptoms resulted from underlying genetic causes, whole-genome sequencing (WGS) was performed on the patient and healthy parents (Supplementary Table 1). Our analysis uncovered two candidate-coding variants in the patient without corresponding genetic alterations in the parents, suggesting these variants arose *de novo*. The first candidate was a mosaic splice site variant *TULP3* (NM_003324.4:c.253+1G>T) present at ∼23% variant allele frequency. *TULP3* variants have not been associated with human disease according to OMIM, and the c.253+1G>T variant does not show constraint for loss-of-function (LoF) variation according to gnomAD (pLI=0), suggesting the splice site variant is not pathogenic. The second variant identified was a *de novo* missense change in *AUTS2* (NM_015570.3:c.1600A>C, p.(Thr534Pro)). This *AUTS2* variant is predicted to be damaging by 16/25 *in silico* tools according to VarSome^27^. Sanger sequencing of the family trio confirmed its *de novo* status (Fig.1d). Although the vast majority of *AUTS2* variants reported as pathogenic or likely pathogenic to the ClinVar database are predicted to cause loss-of-function changes, five distinct missense variants (including c.1600A>C identified in our patient) have been reported within amino acid residues 529-535 (Fig.1e). Sequence alignment of vertebrate amino acid sequences within the H1 region show that this portion of AUTS2 is significantly conserved, suggesting functional importance (Fig.1f). Since few missense variants in *AUTS2* have been reported to cause disease and given its expanding role in a range of neurological disorders, we sought to investigate the underlying pathogenic mechanism of this p.Thr534Pro variant.

### Expression of AUTS2 in excitatory neurons and progenitor cell types

To investigate which cell types may be affected by the *AUTS2*^*T534P*^ variant during corticogenesis, we analyzed a published scRNA-seq CO and fetal brain dataset to evaluate *AUTS2* expression at cell-type specific resolution^19^. These data indicate *AUTS2* is expressed in a diversity of cell populations within the developing fetal brain and COs with notable expression in excitatory neurons, radial glia, and intermediate progenitors (Supplementary Fig. 1). Given the expression of *AUTS2* in these progenitor cell types and the highly dynamic developmental program of NPC proliferation and differentiation during corticogenesis, we sought to investigate its pathophysiology in COs, which are an emerging model system that recapitulate morphological and early neurodevelopmental features of the human brain.

### *AUTS2* patient COs exhibit patient-specific microcephaly and show proliferative deficits in NPCs

To investigate the functional role of the *AUTS2*^*T534P*^ variant during human cortical development, we reprogrammed peripheral blood monocytes from the *AUTS2*^*T534P*^ patient and her parent as a control into human induced pluripotent stem cells (hiPSCs) using Sendai virus-based delivery of Yamanaka transcription factors (Supplementary Fig. 2). hiPSC colonies emerged within seven days post-infection showing tight cell junctions, distinct cell borders, and alkaline phosphatase activity, which are characteristic features of human pluripotent stem cells (Supplementary Fig. 2a,b). We then evaluated the chromosomal integrity and pluripotency of the hiPSCs by karyotyping (Supplementary Fig. 2c) and immunohistochemical analysis of key pluripotency markers, including OCT3/4, SSEA4, NANOG and LIN28 (Supplementary Fig. 2d). Additionally, we confirmed the heterozygous *AUTS2*^*T534P*^ variant was present in the *AUTS2*^*T534P*^ patient line as demonstrated by Sanger sequencing (Fig. 1d). We then used these hiPSC lines to generate three-dimensional COs with our undirected differentiation protocol^28^. Our overall approach for modeling the *AUTS2*^*T534P*^ variant in COs is summarized in Figure 2a.

**Figure 2.**
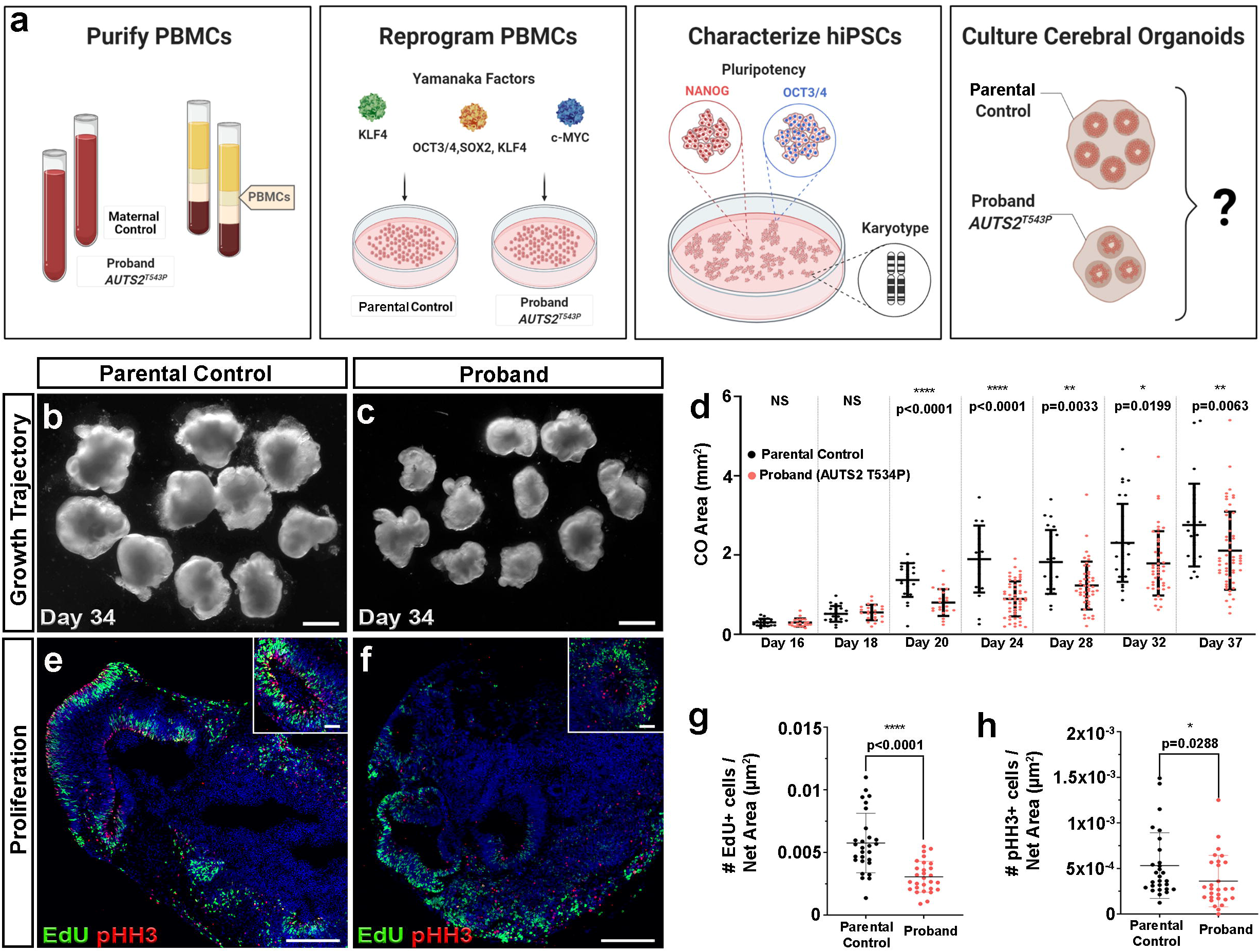
*AUTS2* patient COs show reduced growth and proliferative deficits. (**A**) Overview of protocol used to generate COs. Briefly, peripheral blood mononuclear cells (PBMCs) were isolated from patient and control blood samples and underwent Sendai virus transduction with Yamanaka factors (OCT3/4, SOX2, KLF4, c-MYC) to produce induced pluripotent stem cells (iPSCs). These cultures were characterized and then used to generate COs through undirected, spontaneous differentiation. (**B-C**) Representative images of parental control COs and proband COs at Day 34 showing a significant growth reduction in proband COs. Scale bar = 1 mm. (**D**) Growth trajectory analysis of parental control and proband COs from days 16-37 in CO development. (**E-F**) Parental control COs show proliferating neural progenitors identified as EdU+ and phospho-Histone H3+ (pHH3; mitosis marker) compared to proband COs, which show a reduction (quantified in **G** and **H**). Scale bar = 50 µm. DAPI (blue) stains nuclei. All data are shown as the mean ± standard deviation (SD). Statistical analyses in panel d were performed using Mann-Whitney U tests and those in panels g and h were performed using one-way ANOVA with Tukey’s multiple comparisons test (n = 4 independent organoids per group and 2 independent experiments performed). Statistically significant differences between parental control and proband COs were observed between groups commencing at day 20. *p ≤ 0.05; **p ≤ 0.01; ***p ≤ 0.001; ****p ≤ 0.0001.

To investigate whether proband COs modeled the patient’s microcephaly, we differentiated proband and parental control hiPSCs into COs and performed a rigorous growth trajectory analysis of COs within a dynamic neurodevelopmental time window of NPC proliferation and differentiation (Fig.1b-d). Proband and parental control COs were indistinguishable by size or gross morphology until day 20 of culture, when proband COs showed a statistically significant reduction in overall growth compared to the parental control from day 20 (Fig.1d). This trend persisted over time (day 20 median cross-sectional area of proband COs was 0.71mm^2^ compared to 1.41mm^2^ in parental control COs) (Fig. 2b-d). By day 28, the median cross-sectional area of proband COs was 1.52mm^2^ in proband COs compared to 2.56mm^2^ in parental control COs (coefficient of variation (CV)=49.01% in proband COs and 44.2% in parental control COs). This trend was also observed across multiple cohorts (n=4), suggesting that the proband COs recapitulate morphological features of the patient’s microcephaly.

We then tested whether proliferative deficits in NPCs within ventricular-like zones (e.g. neural rosettes) underly the growth deficits observed in proband COs. To perform this analysis, we pulse-labeled proliferating cells in proband and control COs with EdU^29^, and performed a quantitative imaging analysis for the M-phase specific marker, phospho-histone H3+ (pHH3) (see Supplementary Table 2 for all antibodies utilized in this study), and EdU+ cells within neural rosettes. Our analysis revealed a statistically significant reduction in the number of EdU+ and pHH3+ NPCs in proband COs compared to the parental control (Fig. 2e-h). These data suggest proband NPCs have deficits in cell cycle control, which may underlie the reduced growth properties of proband COs.

### Proband COs exhibit increased asymmetric cellular divisions and ciliary defects

Since cell cycle control and cellular division is tightly coupled within the VZ during human corticogenesis, we investigated the dividing properties of apical progenitors along VZ-like zones in proband COs. During early human brain development, NPCs self-renew through symmetrical divisions along the ventricular surface to sufficiently expand the pool of NPCs allowing for subsequent neurogenesis^30-32^. After this rapid symmetrical expansion of NPCs, they undergo a gradual shift towards dividing asymmetrically to form a daughter NPC and either an intermediate progenitor cell (IPC) or a neuron. Compared to rodent brain development, human brains undergo more extensive symmetrical divisions of apical progenitors and increased basal progenitor proliferation within the oSVZ resulting in a larger frontal cortex^30-32^, a key difference which may explain why mutations causing microcephaly in humans are not adequately modeled in the mouse. Several studies have suggested primary microcephaly results in the loss of cortical expansion caused by premature neuronal differentiation due to the depletion of dividing progenitors^21,22,33^. This process occurs when progenitors prematurely undergo asymmetric divisions to generate neurons instead of additional progenitors at an early stage of neurodevelopment. (Fig. 3a).

**Figure 3.**
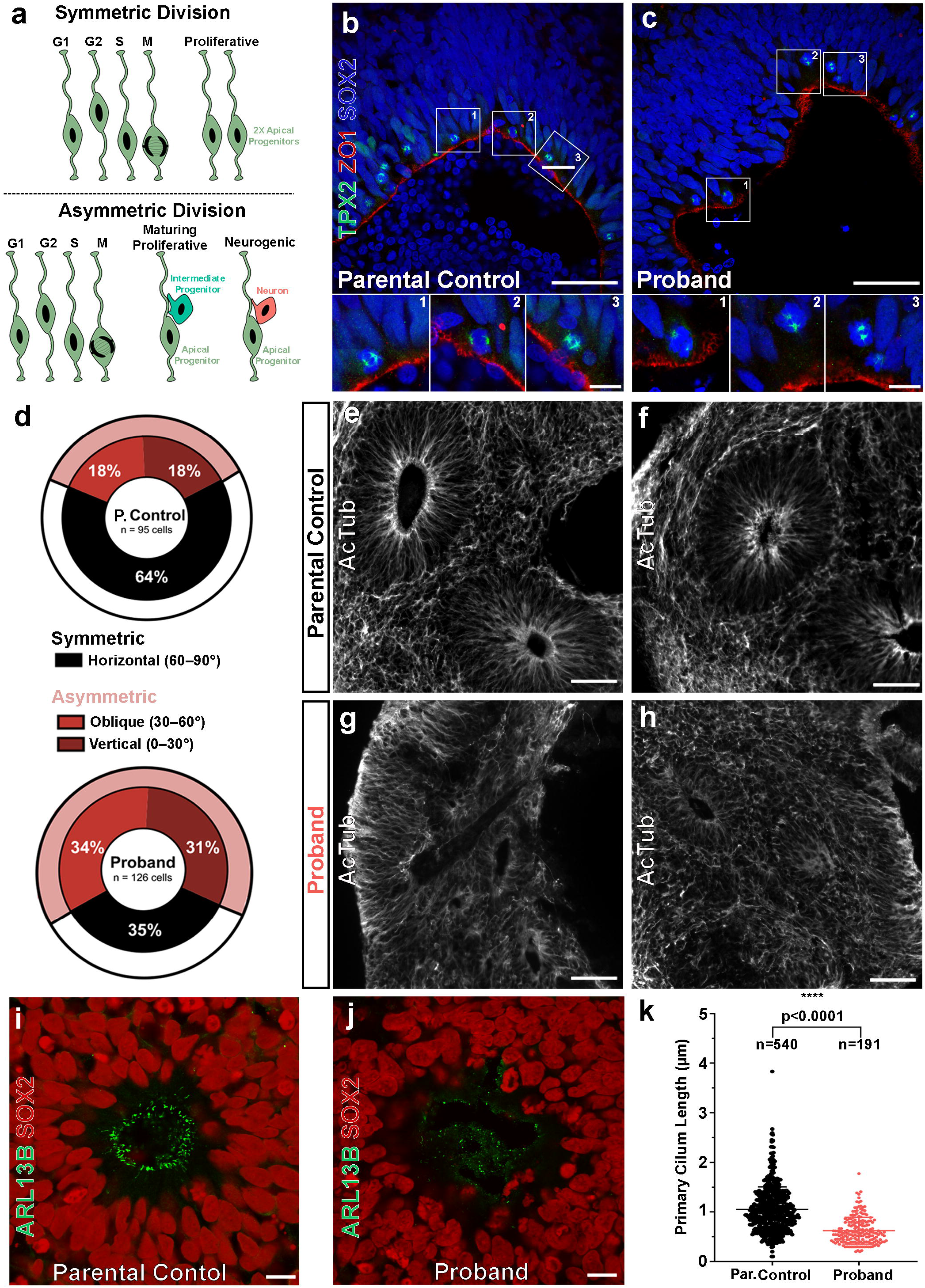
*AUTS2* patient COs show functional and molecular deficits in neural progenitor cells. (**A**) The fate of neural progenitor daughter cells correlates with their mitotic division class. Symmetric divisions (top) give rise to two daughter apical progenitors whereas asymmetric divisions (bottom) give rise to a single apical progenitor and one mature cell type (intermediate progenitor or neuron). (**B-C**) Representative mitotic divisions at the apical surface of parental control and proband rosettes of COs. The division class is determined by the acute angle, θ_a_, that forms between the cleavage plane (defined by TPX2+ mitotic poles, green) and the apical surface (marked by ZO1, red). Cell nuclei visualized with DAPI (blue). Scale bar = 50 µm and 10 µm for lower and higher magnification images, respectively. (**D**) Asymmetric oblique and vertical divisions are overrepresented in proband COs (65%) compared to parental control COs (36%). **(E-F)** Acetylated tubulin (AcTub) staining reveals normal rosette microtubular organization within parental control COs, but severely disrupted organization in proband COs (**G-H**). Scale bar = 250 µm. **(I)** SOX2+ neural progenitors (red) within rosettes of parental control COs form robust, uniform ARLB13B+ cilia (green) at the apical surface, whereas those within rosettes of proband COs show shortened and irregular arrangement (**J**). Scale bar = 10 µm. **(K)** Proband cilia show a statistically significant reduction in length compared to parental control cilia. All data are shown as the mean ± SD. Statistical analysis of cilia quantification data was performed using an unpaired t-test (n =540 parental control cilia and n=191 proband cilia quantified across 4 independent organoids per group and 1 independent experiment performed). ****p ≤ 0.0001.

To test whether a similar mechanism of premature asymmetric divisions occurred in proband NPCs, we measured the division angles of SOX2+ apical progenitors at the VZ within our COs using the mitotic spindle marker TPX2 and apical zone marker ZO1 (Fig. 3b-c). Approximately one third of divisions within parental control COs were asymmetric (vertical: 18%, oblique: 18%, horizontal: 64%; n=95 cells) whereas nearly two-thirds of divisions in the proband *AUTS2*^*T534P*^ COs were asymmetric (vertical: 31%, oblique: 34%, horizontal: 35%; n=126 cells). This approximate two-fold increase in asymmetric divisions within proband COs indicates progenitors may be undergoing premature neuronal differentiation. Taken together, the overall decrease in proliferation and increase in neurogenic, asymmetrical divisions within proband COs suggests that the *AUTS2*^*T534P*^ variant adversely affects NPC cell cycle dynamics and/or cell fate determination.

We next analyzed the primary cilium within neural rosettes as this structure is critical in establishing NPC columnar organization within the VZ and is directly involved in cell cycle kinetics of various cell types, including NPCs. Several microcephalic phenotypes have been linked to disruptions in primary cilium dynamics^22,34-36^. We found VZ-like structures within proband COs that lacked NPC columnar organization and uniform polarity. Specifically, they exhibited disrupted microtubule networks within rosettes, as shown by their irregular acetylated tubulin immunoreactivity (Fig. 3g,h,j) compared to controls (Fig. 3e-f, i). Additionally, we observed a statistically significant reduction in the length of proband ARLB13B+ cilia compared to controls (Fig. 3k). Collectively, these results suggest the proband COs exhibit deficits in ciliary properties and organization with a concomitant loss of NPC polarity within CO rosettes. Since primary cilium formation and resorption is imperative to the progression of the cell cycle, the loss of NPC polarity in proband COs provides further evidence that the *AUTS2*^*T534P*^ variant impairs their proliferative capacity.

Given the established role of AUTS2 in neuronal migration^6^, we then evaluated whether there were any overt abnormalities in progenitor cell migration by analyzing the distribution of TBR2+ intermediate progenitor cells (IPCs) surrounding neural rosettes in proband COs. We did not observe any marked differences in the distribution of TBR2+ IPCs demarcating neural rosettes within both proband and control COs (Supplementary Fig. 3a). Further, no IPCs were detected within proband or control rosettes as expected, suggesting the *AUTS2*^*T534P*^ variant may not substantially affect progenitor migration. To investigate AUTS2 expression in proband COs, we observed both cytoplasmic and nuclear expression of AUTS2 in NPCs, similar to the parental control, and verified AUTS2 staining specificity using a blocking peptide directed against the AUTS2 antibody utilized. These data suggest the *AUTS2*^*T534P*^ variant does not alter normal AUTS2 expression patterns (Supplementary Fig. 3b). We also observed that a subset of IPCs were both AUTS2+ and TBR2+ in both proband and control COs, which is consistent with our synthetic scRNA-seq analysis (Supplementary Fig. 1).

### Growth area and NPC proliferative deficits in proband COs are rescued by gene correction

To investigate whether the phenotypes we observed in proband COs were caused by the *AUTS2*^*T534P*^ variant, we gene corrected this alteration to the wild-type sequence using CRISPR-Cas9 homology directed repair (Fig. 4a). A small guide (sg) RNA (20 base pairs) targeting the *AUTS2* variant region was designed that contained <2 base pair matches to other sequences in the genome to ensure precise gene targeting. Additionally, a silent edit was inserted in the single-stranded oligo donor (ssODN) sequence to disrupt repeated sgRNA binding and subsequent re-cutting by Cas9 after successful editing (Fig. 4b). sgRNA and ssODN sequences are provided in Supplementary Table 3. Synthego’s Inference of CRISPR Edits (ICE) software tool was then used to measure the frequency of successful gene editing. We then generated single iPSC clones, screened ∼100 of them using Sanger sequencing, and confirmed three clones showed both successful gene correction (c.1600C>A) and introduction of the silent gene edit (c.1608G>A). Next, we verified pluripotency markers using immunofluorescence analysis and chromosomal stability using karyotype analysis in the GC hiPSC line prior to generating COs (Supplementary Fig. 2b-d).

**Figure 4.**
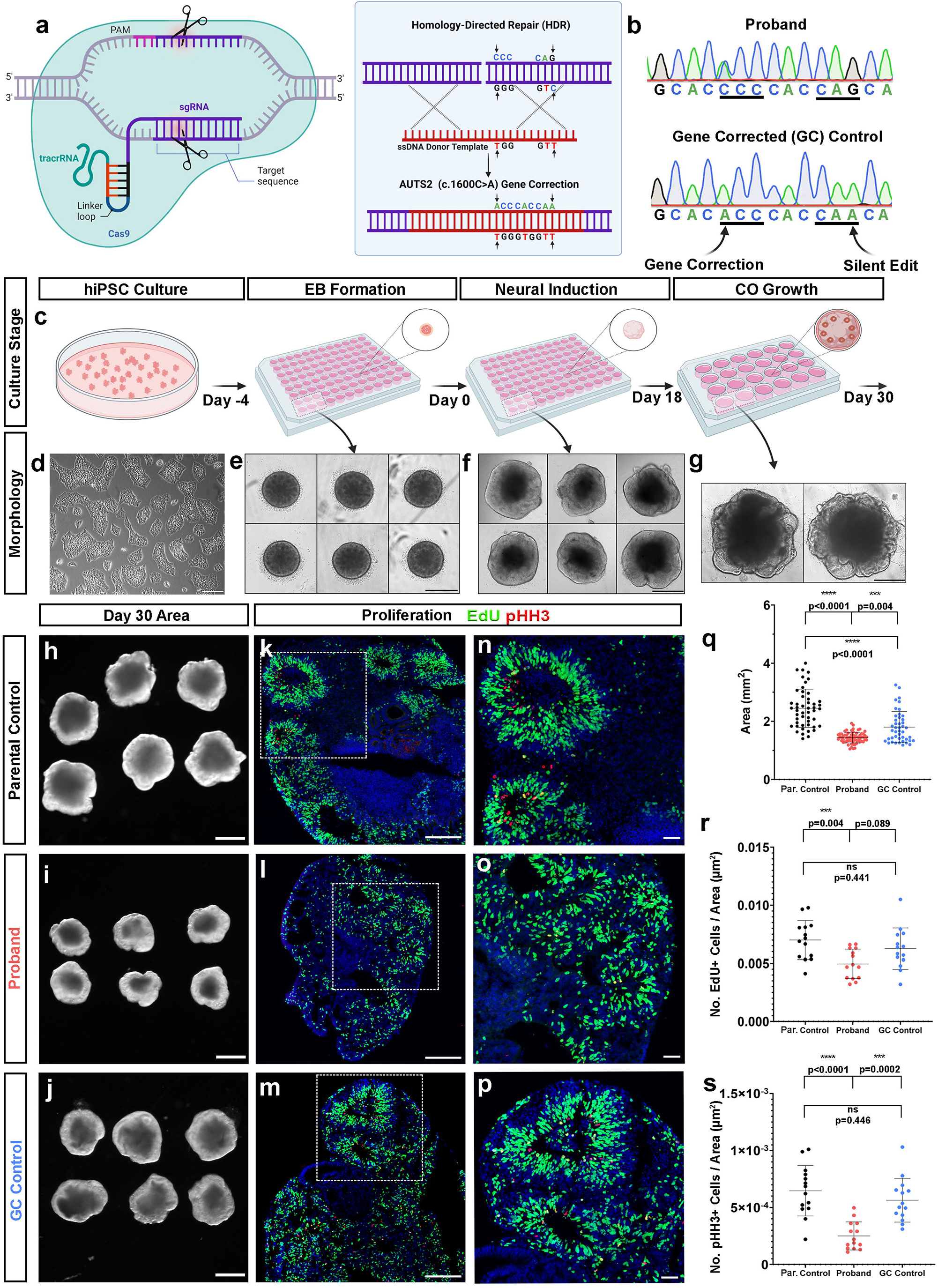
*AUTS2* gene editing with CRISPR-Cas9 rescues proband COs phenotypes. (**A**) CRISPR-Cas9-mediated homology directed repair gene correction strategy (**B**) Chromatogram generated by Sanger sequencing of affected patient (*AUTS2* c.1600A>C, proband) and GC hiPSC line. First arrow denotes the C>A base pair change at c.1600 and the second arrow denotes the silent gene edit (G>A) at c.1608. (**C-G**) Schematic showing major steps in an optimized protocol to generate reproducible COs from hiPSCs with representative culture phase contrast microscopic images below each step. Scale bar = 200 µm and 500 µm for **(D)** and **(E-G)**, respectively. (**H**,**I**,**J**) Representative images of parental control, proband, and GC control COs at Day 30, cross-sectional area of each CO group quantified in (**Q**). (**K**,**L**,**M**) Decreased percentage of EdU+ and phospho-Histone H3+ (pHH3; red, mitosis marker) progenitors in proband CO rosettes compared to parental and GC controls. Scale bar = 200 µm. Magnified in (**N**), (**O**) and (**P**), scale bar = 50 µm; quantified in (**R**) and (**S**). All data are shown as the mean ± SD. Statistical analyses were performed using one-way ANOVA with Tukey’s multiple comparisons test (n = 14 rosettes quantified across a minimum of 4 independent organoids per group and 1 independent experiment performed). ***p ≤ 0.001; ****p ≤ 0.0001; ns=not significant.

We then generated COs using an optimized protocol to enhance organoid reproducibility in our cultures^37,38^ and to test whether GC of the *AUTS2* variant rescued the microcephalic phenotype we observed in proband COs (Fig. 4c-g). This protocol obviates the requirement to encapsulate neuroepithelial bodies and generates morphologically reproducible and robust COs compared to prior protocols^37,38^. Consistent with our previous studies, proband COs generated by this method showed a reduction in growth compared to parental control COs (day 30 median cross-sectional area of proband COs was 1.44 mm^2^ compared to 2.43 mm^2^ in parental control COs) (Fig. 4h,i,q). In contrast, our GC COs showed a statistically significant increase in cross sectional area compared to proband COs, suggesting phenotypic rescue and that the *AUTS2*^*T534P*^ variant contributes to organoid growth deficits (day 30 median cross-sectional area of GC control COs was 1.68 mm^2^, 75^th^ percentile cross-sectional areas were 1.55 mm^2^ in proband COs compared to 2.11 mm^2^ in GC control COs and 2.81 mm^2^ in parental control COs (Fig. 4i,j,q). However, no statistically significant difference in cross-sectional area was observed between the parental control and GC control COs. This finding may suggest the gene correction results in a partial rescue and/or highlights the variability in growth properties of COs arising from intrinsic differences between hiPSC lines (Fig. 4r). The latter scenario underscores the value of performing these types of analyses with a GC hiPSC line. Our optimized CO generation protocol resulted in a significant reduction of the CV in CO area measurements across groups compared to our earlier CO area growth analysis (CV=12.78% in proband COs, 26.86% in parental control COs, and 29.77% in GC control COs), which suggests this improved CO generation protocol lowers organoid-to-organoid variability.

Next, we tested whether NPC proliferative deficits in proband COs were rescued by the *AUTS2* variant gene correction. Similar to our prior experiment, we pulse-labeled proliferating cells in COs with EdU and then performed a quantitative imaging analysis for pHH3+ and EdU+ cells within neural rosettes across all organoid groups (Fig. 4k-p). Although an increased trend in the number of EdU+ was observed in the GC control compared to proband COs, this increase did not reach statistical significance (Fig. 4r). However, a statistically significant increase in the number of pHH3+ NPCs was observed in the GC control compared to proband NPCs (Fig. 4s). In addition, no significant difference was observed between the parental control and the GC control, suggesting phenotypic rescue, and thus providing further evidence that the *AUTS2*^*T534P*^ variant contributes to deficits in NPC proliferation (Fig. 4s).

### Single-cell RNA sequencing reveals a susceptible population of NPCs in proband COs that is rescued by gene correction

To determine cell-type specific transcriptomic signatures underlying the proband COs microcephalic phenotype, we performed scRNA-seq analysis in day 30 proband COs (n=8 organoids pooled per group; Fig. 5). We performed unsupervised clustering on gene expression profiles from an integrated dataset of 35,633 cells and identified ten composite clusters using canonical marker genes (Supplementary Fig. 4). Since the scope of our study primarily concerned NPCs and differentiated immature neurons, we subclustered them for further analysis. This sub-clustering resulted in five cell types from an integrated dataset of 17,752 cells, which were comprised of *EOMES*+ intermediate progenitor cells (IPCs), *STMN2*+/*GAP43*+/*TBR1*+ immature neurons, and three classes of *SOX2+* neural progenitor cells (NPCs) (Fig. 5a,b). Upon initial analysis, we observed a drastic reduction in the relative proportion of Type 2 NPCs in proband COs compared to both controls (Fig. 5c-h). Strikingly, Type 2 NPCs constituted approximately 22% of cells within parental control COs, but only 0.25% in proband COs. This underrepresented NPC population was significantly rescued in GC control COs showing a 22-fold increase of Type 2 NPCs (5.5% of total) compared to proband COs, suggesting the reduction of this cell type is related to the *AUTS2*^*T534P*^ variant. We also observed a two-fold increase in the percentage of immature neurons in proband COs (29%) compared to parental control COs (14%), which supports our earlier results showing increased asymmetrical divisions and subsequent premature neuronal differentiation in proband COs. The GC control also showed a decrease in the percentage of immature neurons compared to proband COs (from 29% in proband COs to 23% in GC control COs), suggesting that gene correction of the *AUTS2* variant partially restores the proper timing of neurogenesis in COs. We also noted a nearly two-fold increase in the percentage of IPCs in proband COs, which constitute 7% of all subclustered cells but only 3-4% in control COs. Analysis of *AUTS2* expression in our sc-RNA seq data showed similar levels across both proband and control groups in NPCs and in immature neurons, suggesting the *AUTS2*^*T534P*^ variant does not affect mRNA stability (Supplementary Fig. 5).

**Figure 5.**
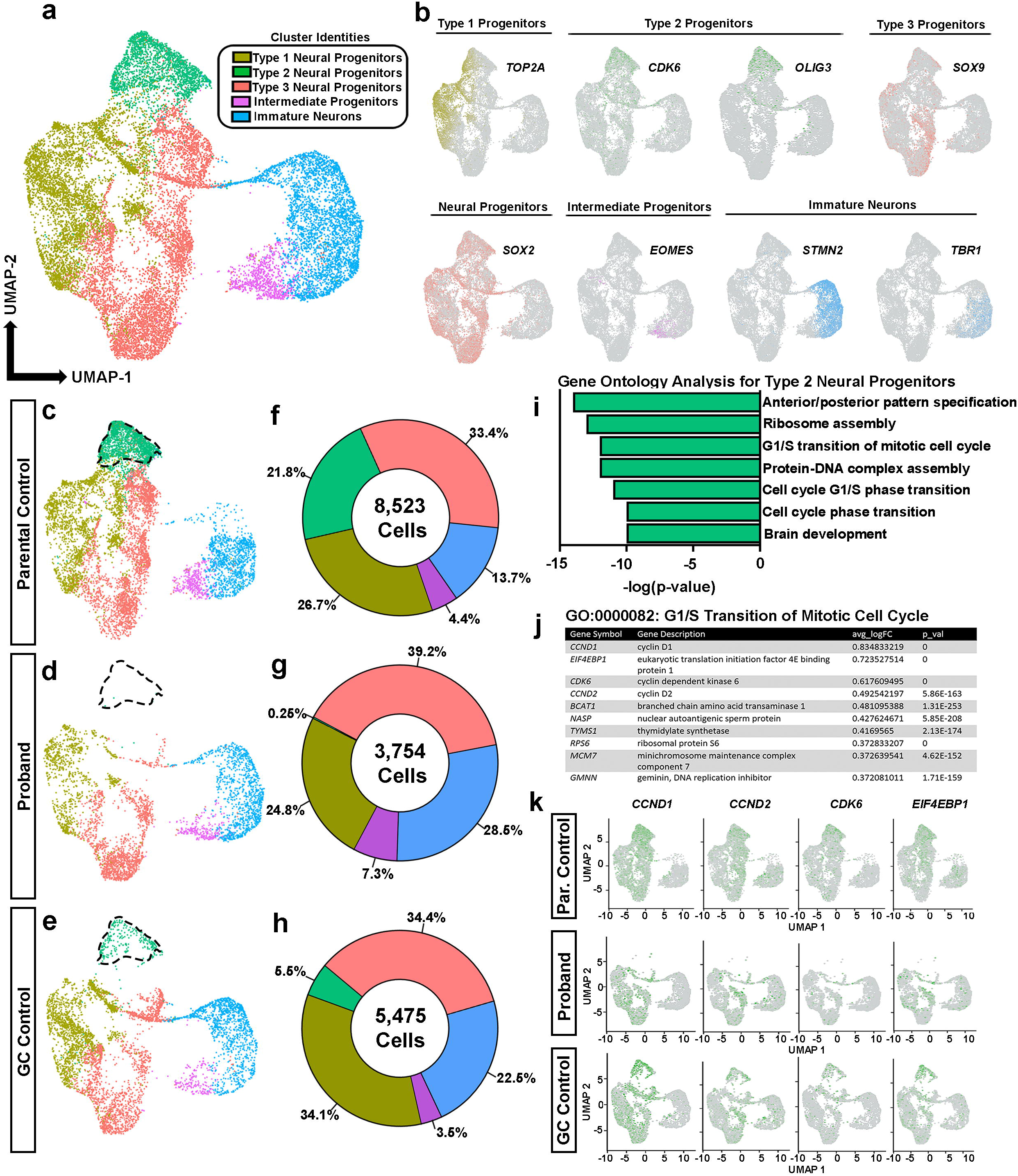
Single-cell RNA sequencing reveals an underrepresented population of proliferative neural progenitor cells in proband COs. (**A**) UMAP plot of key cell types organized into five major clusters: Intermediate progenitors, immature neurons, and three classes of neural progenitor cells containing 17,752 cells. (**B**) Select canonical markers used to determine cluster identities. *MKI67* and *TOP2A* show enriched expression within Type 1 neural progenitor cells, *CDK6* shows enriched expression in Type 2 neural progenitor cells and *SOX9* shows enriched expression in Type 3 neural progenitor cells; *SOX2* is a pan-progenitor marker; *EOMES* (*TBR2*) labels Intermediate Progenitors; *STMN2* and *TBR1* label Immature Neurons. (**C-E**) UMAP plots of cells from parental control COs (8,523 cells), proband COs (3,754 cells) and GC control COs (5,475 cells). Type 2 neural progenitors are outlined in each plot to highlight their underrepresentation in proband COs. (**F-H**) Percentages of cell types per group. (**I**) GO terms identified in Type 2 neural progenitors enriched for genes associated with G1/S cell cycle phase transition. (source: Metascape) (**J**) Differentially expressed genes and associated information identified in the G1/S transition of the mitotic cell cycle gene ontology. (**K**) Feature plots showing expression of G1/S cell cycle genes in proband and GC control COs.

Next, we investigated gene expression signatures enriched in Type 1 and Type 3 NPCs. Type 1 NPCs were enriched for G2/M phase gene expression signatures such as *MKI67* and *TOP2A* and Type 3 NPCs were enriched for *FABP7* and *SOX9*, transcription factors which are critical for NPC proliferation and promote neuronal and glial fate specification^39-42^ (Fig. 5b, Supplementary Fig. 4). Type 1 NPCs were comprised of similar percentages within proband (24.8%) and parental control COs (26.7%) and were elevated approximately by 10% in GC control COs. Type 3 NPCs showed an approximate 5% increase in proband COs compared to both control COs. The significance of these differences is unknown and warrants future investigation.

To determine specific biological processes and molecular functions that characterize Type 2 NPCs, we performed a gene ontology analysis from the top enriched gene expression signatures identified in Type 2 NPCs (Fig. 5i). Type 2 NPCs were enriched for the GO terms: anterior/posterior pattern specification (GO:0009952), ribosome assembly (GO:0042255), G1/S transition of mitotic cell cycle (GO:0000082), protein-DNA complex assembly (GO:0071824), and others (Fig. 5i). Further analysis of gene signatures identified within the anterior/posterior pattern specification gene ontology revealed genes associated with midbrain and hindbrain specification (e.g. *OLIG3, HOXA2*) (Supplementary Fig. 6). Thus, AUTS2 may play a critical role in specifying these cells fates and may explain why AUTS2 syndrome patients display cerebellar hypoplasia. Additionally, the Type 2 NPCs were enriched for *CCND1, EIF4EBP1, CDK6, CCND2*, and other gene expression signatures, all of which contribute to the G1-S transition phase of the cell cycle (Fig. 5j). Expression analysis of these cell cycle genes showed significant reductions in proband COs, which were restored in GC COs (Fig. 5k). Collectively, these findings suggest that the *AUTS2*^*T524P*^ variant in proband Cos leads to a selective loss of proliferating progenitors, which is consistent with a reduced percentage of EdU+ and pHH3+ progenitors and consequential microcephalic phenotype in proband COs compared to controls (Fig. 2e-h,4h-t). These results suggest AUTS2 plays an important role in the progression of the cell cycle within NPCs, which is critical for proper timing of human corticogenesis.

**Figure 6.**
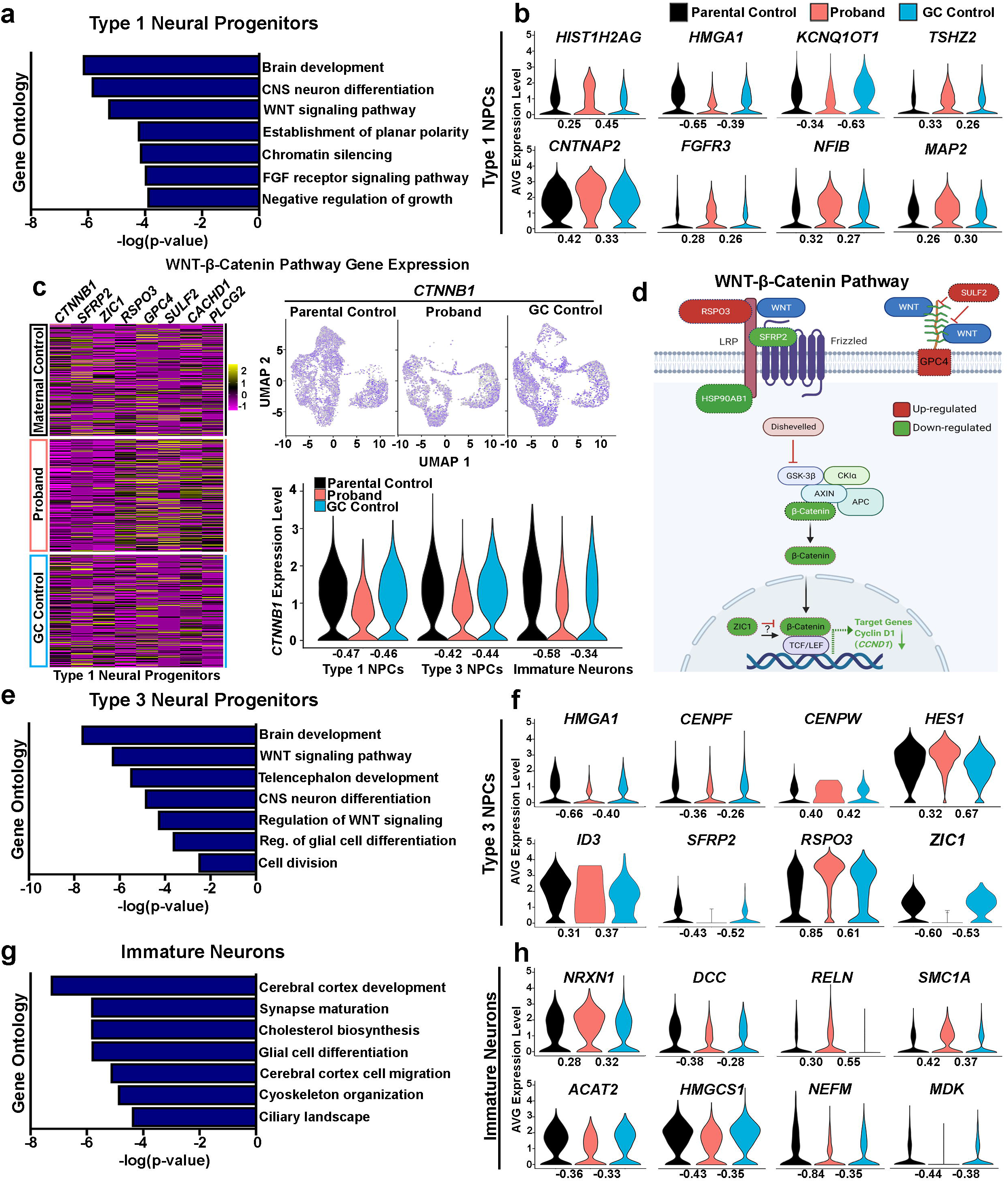
Deficits in WNT-β Catenin pathway gene expression in *AUTS2* patient COs. (**A**) GO terms identified in Type 1 neural progenitors enriched for genes associated with the WNT-β Catenin signaling among others. (**B**) Violin plots of DEGs in Type 1 neural progenitors shows chromatin modifying genes: *HIST1H2AG, KCNQ1OT1*, and *HMGA1*; transcriptional regulator, *TSHZ2*, which has been identified as an AUTS2 target gene; and genes associated with neural and glial differentiation: *FGFR3, NFIB, MAP2;* and cell adhesion: *CNTNAP2*, also identified as an AUTS2 target gene. **(C)** Heat maps of Type 1 neural progenitor DEGs in proband and control COs show alterations in gene expression associated with WNT-β Catenin signaling. Feature and violin plots of *CTNNB1* gene expression in Type 1 and 3 neural progenitors, and immature neurons shows reduced expression in proband COs compared to controls. **(D)** Schematic showing the WNT-β Catenin pathway and color-coded Type 1 neural progenitor DEGs (Red=up-regulated and green=down-regulated). **(E)** GO terms identified in Type 3 neural progenitors enriched for genes associated with the canonical WNT signaling pathway, regulation of glial cell differentiation, and others. **(F)** Violin plots of DEGs in Type 3 neural progenitors show NOTCH signaling transcription factor, *HES1, and ID3*, which regulate glial differentiation; genes regulating cellular division: *CENPF* and *CENPW*; chromatin modifying gene, *HMGA1*; and WNT signaling regulators: *RSPO3, SFRP2*, and *ZIC1* **(G)** GO terms identified in immature neurons enriched for genes associated with synapse maturation, cholesterol metabolism, cytoskeletal organization, and others. **(H)** Violin plots of DEGs in immature neurons shows genes associated with synapse maturation: *NEFM, RELN*, and *NRXN1*; cholesterol biosynthesis: *ACAT2* and *HMGCS1*; and genes associated with cytoskeletal organization: *SMC1A* and *NEFM;* and neurite morphogenesis: *DCC* and *MDK*. Of note, *NRXN1* (AUTS2 target gene), *DCC, RELN* (AUTS2 target gene), and *SMC1A* are also designated SFARI genes implicated in autism spectrum disorder and intellectual disability. Log_2_ fold changes (FC) are shown under each violin plot and denote FC relative to either the parental control or gene corrected control.

### Proband NPCs show dysregulated gene expression associated with WNT-β-catenin signaling, chromatin modification, and gliogenesis

To further investigate underlying molecular mechanisms affecting NPC proliferative deficits in *AUTS2* patient COs, we performed gene ontology analyses from genes identified within the Type 1 and 3 NPC clusters that were differentially expressed in comparison to both parental and GC control COs. In Type 1 NPC, we observed differentially expressed genes (DEGs) that were enriched for the GO terms: brain development (GO:0007420), CNS neuron differentiation (GO:0021953), WNT signaling pathway (GO:0016055), chromatin silencing (GO:0006342), and others (Fig. 6a). Of note, the chromatin modifying gene, *HMGA1*, showed reduced expression in proband COs (Fig. 6b), which has been shown to play an important role in controlling the neurogenic potential of NPCs and the developmental timing of gliogenesis^43,44^. Other chromatin modifying genes, *HIST1H2AG* and *KCNQ1OT1*, were also dysregulated. Our analysis also identified increased expression levels of *MAP2*, which is consistent with our earlier finding that proband NPCs are undergoing premature neuronal differentiation. *NFIB* and *FGFR3* expression levels were also induced in proband Type 1 NPCs, suggesting these progenitors show a propensity towards glial cell differentiation. Interestingly, we observed increased expression of the homeobox transcription factor, *TSHZ2*, and the neural cell adhesion gene, *CNTNAP2*, both of which have been previously identified as AUTS2 target genes in mouse studies^45^ (Fig. 6b). We then focused our analysis on those DEGs associated with WNT signaling as numerous studies have underscored its critical role in regulating NPC proliferation and differentiation within the ventricular zone^46-48^. *CTNNB1*, which is the major signal transducer of WNT signaling, was significantly reduced in proband COs within both Type 1 and 3 NPCs and immature neurons. We also observed alterations in the expression of multiple genes in proband COs compared to control COs, which function in regulating WNT signaling activity such as *RSPO3, SFRP2, GPC4*, and *SULF2* (Fig. 6c,d). Cyclin D1 (encoded by the *CCND1* gene) is a well-characterized target for WNT-β-catenin signaling, which promotes G1/S phase transition of the cell cycle^49,50^, was reduced in proband NPCs compared to controls (Fig. 5k). Collectively, our data suggests that proband NPCs show deficits in WNT-β-catenin signaling, which may underly their proliferative deficits and impairment to transition through the G1/S phase of the cell cycle.

We then performed the same type of GO analysis for Type 3 NPCs and observed DEGs that were enriched for the GO terms: brain development (GO:0007420), WNT signaling pathway (GO:0016055), regulation of glial cell differentiation (GO:0045685), cell division (GO:0051301), and others (Fig. 6e). Similar to Type 1 NPCs, Type 3 NPCs showed reduced expression of *HMGA1* and dysregulated expression of the WNT signaling regulators, *RSPO3, SFRP2*, and *ZIC1*. In addition, we observed dysregulated expression of the centromere binding genes, *CENPF* and *CENPW*, which may contribute to the polarity and mitotic deficits in proband NPCs compared to controls. Increased expression of the *HES1* and *ID3* genes were also observed in proband NPCs. Previous studies have suggested that HES1 and ID3 promote the specification of neural precursors towards an astrocyte fate^51,52^, which suggests proband NPCs may have an altered propensity to differentiate into glial identities.

Next, we examined the DEGs identified in immature neurons from our sc-RNA seq data. Mouse studies have underscored a critical role for AUTS2 in controlling neuronal gene expression and specifying neuronal cell fates, however few studies have examined the role of AUTS2 in human corticogenesis and its biological link to intellectual disability observed in AUTS2 syndrome patients. Similar as described above, we performed a GO analysis on DEGs from immature neurons and identified the following enriched biological processes: synapse maturation (GO:0060074), cholesterol biosynthesis (R-HSA-191273), glial cell differentiation (GO:0010001), and cytoskeleton organization (GO:0045104), among others (Fig. 6g). We observed dysregulated expression in the synaptic maturation genes, *NEFM, RELN, NRXN1*, and reduced expression of genes controlling cholesterol metabolism such as *ACAT2* and *HMGCS1* (Fig. 6h). Other altered gene expression levels were observed in genes regulating cytoskeletal organization: *SMC1A* and *NEFM*; and neurite morphogenesis: *DCC* and *MDK*. Interestingly, *Nrxn1* and *Reln* were previously identified as AUTS2 target genes in mouse studies using ChIP Seq^45^. In addition, *NRXN1, DCC, RELN*, and *SMC1A* have all been implicated in ASD and ID and are designated Simons Foundation Autism Research Initiative (SFARI) genes^53-57^. Further studies are warranted to understand the impact of these altered gene expression patterns on synaptic maturation and activity during neurodevelopment using *in vitro* and *in vivo* models of *AUTS2* deficiency and whether they are linked to cognitive deficits in ASD/ID patients.

## DISCUSSION

This study provides novel molecular insights into AUTS2 function within NPCs, which may underly the neurological manifestations of microcephaly and syndromic intellectual disability observed in AUTS2 syndrome patients. Previous animal studies have identified a role for AUTS2 in transcriptional activation, RNA metabolism, and cytoskeletal regulation in excitatory neurons. However, these animal models of AUTS2 deficiency do not develop reductions in cortical volume (i.e. microcephaly) and therefore may not adequately recapitulate disease mechanisms underlying AUTS2 syndrome within the cerebral cortex.

Here we present a patient with a *de novo* pathogenic *AUTS2*^*T534P*^ missense variant who presents with AUTS2 syndrome. We established a CO model of this patient to investigate the human-specific pathogenesis of *AUTS2*^*T543P*^ in the early developing brain. Our results indicate that AUTS2 deficiency leads to a microcephalic phenotype, which dysregulates cell cycle dynamics within NPCs, leading to a subsequent reduction in proliferation, ciliary defects and the selective loss of progenitors enriched for gene expression associated with G1/S cell cycle transition. Further, scRNA-seq analysis revealed deficits in WNT-β Catenin signaling in NPCs, which may underly their proliferative deficits. Multiple studies have underscored the critical role of β-Catenin within NPCs during neurodevelopment. For example, conditional deletion of β-Catenin in mouse cortical NPCs led to reduced cell proliferation and disruptions in the organization of the neuroepithelium^47^. Conversely, overexpression of a stabilized form of β-Catenin in NPCs increased cell proliferation^46^. Additionally, focal depletion of β-Catenin within NPCs of the mouse ventricular zone caused premature cell cycle exit and neuronal differentiation^48^. However, it is unclear how AUTS2 deficiency is related to deficits in WNT-β Catenin signaling, although a few studies have suggested WNT-β Catenin signaling and AUTS2 share common downstream targets. For example, the WNT target gene, *Cachd1*, was recently identified as an RNA transcript that can be bound and regulated by AUTS2^14^. Other shared AUTS2 targets identified using ChIP Seq^45^ and targets of Wnt signaling using RNAseq^58^ include *Fzd1* and *Nfia*. It is also plausible that altered expression of chromatin modifying genes observed in proband NPCs may dysregulate downstream WNT target gene expression. Interestingly, *CTNNB1* gene mutations have been associated with dysmorphic features, microcephaly, and intellectual disability in patients^59,60^, clinical symptoms all of which have been observed in AUTS2 syndrome patients. Further investigation is required to determine whether overexpression of *CTNNB1* in *AUTS2*^*T543P*^ COs can rescue NPC proliferative deficits.

More recent work by Liu *et al* demonstrated the HX domains of AUTS2, including the HQ-rich domain within exon 9, are critical for the previously characterized AUTS2-P300 complex^61^. Variants that fall within this HQ-rich domain are associated with neurological symptoms such as microcephaly and intellectual disability–a strikingly similar clinical presentation to Rubinstein-Taybi syndrome, which is caused by pathogenic variants in *CREBBP/P300*. In this study, the AUTS2^T534P^ variant is deficient in recruiting P300 to the PRC1.5-AUTS2 complex, leading to transcriptional dysregulation of downstream target genes. Although this work provides important insight to the mechanism by which the AUTS2^T534P^ variant alters PRC1.5-AUTS2 activity, our study provides novel molecular insights into how this variant leads to pathogenesis in the developing human brain.

One of the challenges associated with CO modeling is variability associated with utilizing different hiPSC lines and organoid-to-organoid variability present within a particular experimental batch. We addressed these challenges by 1) adapting a modified CO protocol to enhance organoid reproducibility, and 2) generating an isogenic, gene-corrected hiPSC line. By gene correcting this variant in proband hiPSCs, we determined the proliferative and molecular deficits observed in proband NPCs were directly linked to the *AUTS2* variant. In GC COs, we observed a statistically significant rescue of overall growth compared to proband COs, although overall growth of GC COs did not match that of parental control COs. However, at the molecular level, GC COs showed a statistically significant rescue in the percentage of proliferating pHH3+ NPCs compared to proband COs. Additionally, sc-RNAseq analysis of GC COs showed a rescue of molecular alterations underlying proliferative deficits within proband NPCs.

Our sc-RNAseq analysis also uncovered an NPC population that was strikingly underrepresented in proband COs. GO analysis revealed these Type 2 NPCs were enriched in gene expression signatures associated with G1/S cell cycle transition in addition to marker genes associated with midbrain and hindbrain specification. The loss of these cell fates in combination with proliferative deficits observed in proband NPCs, most likely contributes to the microcephalic phenotype observed in proband COs. Interestingly, the AUTS2 syndrome patient described here also displays cerebellar hypoplasia (Fig. 1b), which may be the result of deficits in hindbrain fate specification. Investigation of the *AUTS2*^*T534P*^ variant using cerebellar organoids would provide greater mechanistic insight into AUTS2 function during cerebellum development.

Future studies are required to expand upon our findings and evaluate the mechanisms by which AUTS2 controls cell cycle progression in early NPCs. Additionally, other disease-causing variants, such as those found within the mutation hotspot within the ninth exon of *AUTS2*, warrant further investigation. Since AUTS2 is a master neuronal transcriptional activator, microcephaly in AUTS2 syndrome patients may arise from dysregulation of multiple downstream target genes. Thus, future studies are required to elucidate how transcriptional activation of *AUTS2* variants is altered in NPCs and differentiated neuronal progeny. Our results show a critical role for AUTS2 in NPC proliferation and neuronal specification during early human cortical development, deficits of which may contribute to the clinical manifestations observed in AUTS2 syndrome patients. In sum, this study highlights the value of COs to advance our understanding of mechanisms underlying AUTS2 syndrome with the ultimate goal of developing therapeutic strategies for patients.

## MATERIALS & METHODS

### Subjects

The proband and both of her parents were enrolled as part of an Institutional Review Board (IRB) approved study (IRB:11-00215: Rare Diseases/Genome Sequencing) within the Steve and Cindy Rasmussen Institute for Genomic Medicine at Nationwide Children’s Hospital. Informed consent was provided for all study participants to partake in this research and to have this work published. Genomic analysis was performed on DNA isolated from either peripheral blood or saliva samples.

### Whole genome sequencing and analysis

Whole-genome sequencing was performed for all three members using an Illumina HiSeq4000 instrument according to manufacturer protocols. Reads were mapped to the GRCh37 reference sequence, and secondary data analysis was performed using Churchill (Kelly et al., 2015), which implements the GATK “best practices” workflow for alignment, variant discovery and genotyping. Variants were called using GATK 4.0.5.1, and the resulting VCF file was annotated with genes, transcripts, function classes, damaging scores, and population allele frequencies using an in-house pipeline built around the SNPeff annotation tool.^62^ Our general approach to variant annotation and prioritization has been previously described.^63^ After removing common variants (MAF>0.01 in the gnomAD v.2.1.1 database), we selected for further analysis all splice site, frameshift, and nonsense variants, as well as missense variants predicted to be damaging by SIFT (score<0.05), Polyphen (score>0.453), GERP (score>2.0), or CADD (Phred score>15). Because of the severe presentation and lack of a significant family history, we prioritized candidate de novo mutations consistent with dominant inheritance, but recessive and X-linked models were also considered.

### CO culture

#### iPSC cell line generation and quality control testing

Peripheral blood mononuclear cells (PBMCs) were reprogrammed to human induced pluripotent stem cells (hiPSCs) using the CytoTune-iPS 2.0 Sendai Reprogramming Kit (ThermoFisher) according to manufacturer’s instructions. All hiPSC lines used in this assay were rigorously tested for pluripotency markers, tested negative for mycoplasma, and underwent short tandem repeat (STR) profiling analysis (LabCorp) to authenticate purity of cell lines. In addition, all hiPSC lines were tested for live alkaline phosphatase activity using the Alkaline Phosphatase Live Stain kit (Thermofisher) according to manufacturer’s instructions.

#### iPSC culture maintenance

All lines were maintained under feeder-free and defined, serum free medium conditions. iPSCs were cultured in either mTeSR (StemCell Technologies) or Essential 8(tm) Medium (ThermoFisher) and passaged on vitronectin-coated tissue culture plates using standard methods.

#### CO generation

All media formulations for CO generation are described in Supplementary Table 4. The initial cohorts of whole brain CO used in Figures 2 and 3 were generated according to our previously described protocol^28^. CO culture media were exchanged every third day. An adapted whole brain CO generation protocol was used for subsequent analyses in Figures 4 and 5 optimized to increase CO reproducibility. In this CO protocol, iPSC lines were thawed from cryopreservation and passaged a minimum of one time and maintained in culture for at least seven days. iPSCs at 65-85% confluency were pre-treated with 5µM Y-27632 ROCK Inhibitor for 1 hour and then processed into single cells with TrypLE dissociation reagent (ThermoFisher). Cells were then resuspended in iPSC media supplemented with 5µM ROCK inhibitor at a concentration of 40 cells/µl. Then, 100µl of the suspension was transferred into each well (4,000 cells/well) of a non-tissue culture u-bottom 96-well plate. The plates were then balanced and spun down at 400g for 4 minutes. Although the embryoid bodies (EBs) began to noticeably form within a few hours of seeding, plates were left undisturbed for 72 hours for optimal EB formation. After EB formed, media and unincorporated cells were aspirated from each well and replaced with 150µl of fresh Neural Induction Medium (NIM). The NIM was replaced every other day until Day 10. On Day 10, EBs were transferred to a larger plate format (10cm^2^ untreated dishes) with Cerebral Organoid Expansion Medium (COEM) + 2% Matrigel and placed on an orbital shaker. Then, 72 hours later, the COEM was removed and replaced with Cerebral Organoid Growth & Differentiation Medium (COGDM) + 1% Matrigel. Organoids received fresh COGDM three times per week for the remainder of their time in culture. All media formulations were prepared according to our previous studies^28^.

### *AUTS2* exon 9 PCR amplification and Sanger DNA sequencing

Polymerase chain reaction (PCR) was performed using the JumpStart REDTaq ReadyMix Reaction Mix (Sigma) with the following experimental conditions: 200ng of genomic DNA, 25µM of primers (*AUTS2* Exon9-Forward primer: 5′-TCTTGCGACAGGAACTGAACA-3’, *AUTS2* Exon9-Reverse primer: 5′-GTGCTCTACTTATCCTCACATTTTGC-3’), and the JumpStart REDTaq ReadyMix. PCR cycling parameters were the following: initial denaturation 2Lminutes at 94° followed by 35 cycles at 94° for thirty seconds, thirty seconds at 60°, one minute at 72°, and a final extension of five minutes at 72°. Agarose gel electrophoresis was used to visualize PCR products and then were excised and extracted using the QIAquick Gel Extraction Kit (Qiagen). Purified PCR products were then processed for Sanger DNA sequencing (Eurofins) using both *AUTS2* Exon9F and *AUTS2* Exon9R primers.

### CRISPR/Cas9 *AUTS2* variant gene correction and iPSC clone screening

The patient hiPSC line harboring the *AUTS2*^*T534P*^ variant was gene corrected in collaboration with Synthego with the specific gene correction and silent edit: *AUTS2*^*P534T*^ (c.1600C>A, c.1608G>A) (Fig. 4b). A small guide (sg) RNA (TGTGCTGGTGCGTGTGCTGG) was designed that contained <2 base pair matches to other sequences in the genome to ensure precise gene targeting. The sgRNA was then complexed with Cas9 to generate a ribonucleoprotein complex and together with the single-stranded oligo DNA donor (Supplementary Table 3) were nucleofected into *AUTS2*^*T534P*^ hiPSCs. Synthego’s Inference of CRISPR Edits (ICE) software tool was then used to measure the frequency of successful gene editing. iPSCs were then dissociated with TrypLE Express (Thermofisher) and seeded at 0.5 cells/well into a 96 well dish and allowed to expand to confluency. Genomic DNAs from single iPSC clones were then isolated and screened using Sanger sequencing to confirm successful gene correction and introduction of the silent gene edit.

### Conservation analysis

The full length AUTS2 isoform sequence (AUTS-isoform 1 on UniProtKB) were selected from UniProtKB database. UniProtKB’s in-built ClustalW (Clustal Omega) alignment tool was used to perform sequence alignment. The following default alignment parameters were used-default transition matrix Gonnet, gap penalty of 6 bits, gap extension of 1 bit. The default alignment algorithm HHAlign (Soding J, 2005.) was used to perform sequence alignment.

### Collation of *AUTS2* exon 9 variants

Exon information of *Homo sapiens AUTS2* full length isoform (Ensmbl ID-ENST00000342771.10) was determined based on UCSC Genome browser annotation. Based on this exon 9 mapping, all unique *AUTS2* exon 9 variants on ClinVar, LOVD (Leiden Open Variant Database) and HGMD were collected and tabulated.

### Immunohistochemistry

#### Fixation

Cerebral organoids (COs) were fixed in 4% Paraformaldehyde (PFA, Electron Microscopy Sciences, 15713)/DPBS (Gibco, 14190-144) at 4°C overnight. The next day, COs were placed on a shaker in PFA solution at RT for 10-15 minutes to finalize fixation. Once entirely fixed, residual PFA was removed with three DPBS washes. COs then underwent a sucrose gradient: first, COs were equilibrated to 10% Sucrose (Sigma Life Sciences, S7903-250G)/1% Antibiotic-Antimycotic (Gibco, 15240-062)/ DPBS solution overnight. Then, COs were transferred to a 30% sucrose/1% Antibiotic-Antimycotic/DPBS solution to equilibrate overnight again.

#### Preparation for Cryosectioning

The COs and the 30% sucrose solution were inverted into a petri dish and transferred to excess optimal cutting temperature (OCT) solution (Sakura Finetek USA Inc, 4583) with a sterile, trimmed transfer pipet. Up to five COs were placed quickly into the mold, minimally rearranged in a grid-like fashion, and immediately frozen in a bath of dry ice pellets and methanol. The COs were stored at - 80°C until sectioned.

#### Cryosectioning

The embedded tissue was removed from -80°C and mounted on a cryostat chuck with dry ice. The mounted tissue was placed in the cryostat (ThermoFisher, 957020) to equilibrate to -14°C for approximately one hour. Then, the tissue was sectioned at 20µm directly onto positively charged glass slides (Fisherbrand, 1255017) in the cryostat. The slides were stored at -30°C until further processing.

#### Staining

Slides containing sections were then thawed at room temperature (RT) for at least twenty minutes, outlined with a hydrophobic maker (Life Technologies, 008899), and re-hydrated in PBS. Slides were then incubated with blocking solution containing 0.1% Triton X-100/10% donkey serum/Tris Buffer Solution (TBS) for one hour at room temperature. Slides were then incubated with primary solution (blocking solution, antibodies per dilutions described in Supplementary Table 2) for 16-24 hours at 4°C. Slides were then rinsed three times with TBS solution. Then, secondary solution (blocking solution, species-appropriate secondary antibodies and DAPI) was added and incubated for 2 hours at RT. Residual secondary antibodies were removed with three TBS washes. Slides were then immediately cover-slipped with Fluoromount-G® (SouthernBiotech, 0100-01).

#### Imaging

Images were captured with confocal microscopy on a scanning laser confocal microscope (Zeiss LSM 800).

### CO phenotype characterization

#### Organoid area quantification

Organoid cross-sectional areas were extracted from phase contrast images using FIJI and analyzed for trends in GraphPad Prism X.

#### Mitotic angle analysis

Angle calculations were determined using fixed organoid slices stained with TPX2, ZO1, and DAPI to label spindle microtubules during mitosis, the ventricular surface, and cell nuclei, respectively. First, the ventricular surface was outlined using the Adobe Photoshop 2020 Curvature Pen Tool with a solid line one pixel in width. Next, the Add Anchor Point Tool was used to ensure the outline closely adhered to the border of the ventricular surface. ZO1 and DAPI were used to describe the ventricular surface. Second, the line of cleavage was similarly drawn between the TPX2 signals of cells caught in mitosis. Third, the line of cleavage was copied and transposed to intersect the ventricular surface outline at its closest point using the Path Selection Tool. Fourth, the outlined images were quantified on ImageJ (FIJI) using the Angle tool to measure the angle of cleavage as defined by the acute angle formed by the transposed line of cleavage and the intersecting portion of the ventricular surface for each cell. Finally, each cell was given a division classification of vertical, oblique, or horizontal according to its angle of division falling between 0-30°, 30-60°, or 60-90°, respectively.

#### EdU and pHH3 analysis

First, rosette boundaries were drawn on a merged image of Edu, pHH3 and DAPI signal using the Polygon Selection Tool in FIJI. Then, EdU+ and pHH3+ cells were then manually quantified, and trends were analyzed in GraphPad Prism X.

#### Cilia length and count analyses

Tissue sections from the parental control and proband lines were stained with ARL13B, SOX2 and DAPI according to our IHC protocol. The sections were then surveyed to locate and collect z projection images of rosettes. The upper and lower boundaries of the z projection were defined by ARL13B signal (i.e. appearance of cilia). A maximum projection was then generated and used in subsequent analyses, where cilia length and count as well as rosette area were determined using the ROI and measurement tools in FIJI (ImageJ v.2.0.0-rc-69/1.52p). All statistical analyses were performed using one-way ANOVA with Tukey’s multiple comparisons test.

### Single-cell RNA sequencing sample processing

Cerebral organoids were pooled in groups of eight (per line) and prepared for single-cell RNA sequencing as previously described^28^. In brief, organoids were dissociated into a single cell suspension using a gentleMACS Octo Dissociator (Miltenyi) with the manufacturer’s “37 °C_ABDK_02” program. Then, 2 mL solution of Accumax (Sigma, A7089) was transferred into a gentleMACS C-tube (Miltenyi, 130-093-237), organoids were placed in this solution and processed using an Octo-Dissociator instrument (Miltenyi). After the program completed, 10mL of DPBS + 0.04% BSA (Sigma, A9418) were added to each C-tube and then sample solution was filtered through a 70μm strainer to remove undissociated tissue and debris. Dissociated and strained cell mixtures were then centrifuged for two minutes at 300g and resuspended in 1mL DPBS + 0.04% BSA. Additional cell filtration steps was performed to remove fine cell debris using a 40 μm Flowmi® cell strainer (Sigma, BAH136800040). We found repeating this filtration step twice yielded a cell suspension devoid of cell debris for all lines. The concentration and viability of each suspension was then manually determined using trypan blue staining and a hemocytometer. Finally, samples were diluted to achieve a concentration of 1,000 cells/μL. Approximately 10,000 cells were used to generate single cell RNA-seq libraries using the 10x chromium single cell 3’ V2 library kit (10X Genomics) according to the manufacturer’s instructions and sequencing was performed on a NovaSeq 6000 System (Illumina).

### Single-cell RNA sequencing data processing and analysis

Raw base call sequencing data were demultiplexed with Cellranger (v6.0.0) mkfastq function and aligned by CellRanger (v6.0.0) count function using default settings and the GRCh38-2020-A reference transcriptome from 10x Genomics. Summary by sample: Parental control – 17,066 cells, 23,427 mean reads per cell, 2,370 median genes per cell, 5,850 median UMI counts per cell; Proband – 10,542 cells, 37,722 mean reads per cell, 3,300 median genes per cell, 9,578 median UMI counts per cell; GC control – 14,124 cells, 27,886 mean reads per cell, 2,780 median genes per cell, 6,556 median UMI counts per cell. The count matrices were then converted to objects in Seurat v3.3.0 ^64,65^ for analysis and to generate visual representations. Standard quality control parameters were applied to each object. In brief, cells with greater than 200 and less than 5,000 unique features or less than 20% mitochondrial reads were retained. The data were log-normalized with a scale factor of 10,000 and the top 2,000 variable features per object were identified. Integration anchors were generated with 30 principal components and used to integrate all samples into a combined object with a total of 35,633 cells. Standard Seurat procedures were applied to perform a principal component analysis and KNN unbiased clustering. The FindNeighbors (PCA reduction, 30 dimensions) and FindClusters (0.5 resolution, 30 dimensions) functions were used to identify 24 unbiased clusters. Cluster identities were determined by characteristic canonical markers^38,66-68^ and 5 unique clusters were identified. Given the scope of the study, we opted to utilize a subset object with 17,752 cells that exclusively contained clusters relevant to our focus on progenitor-to-neuron differentiation. This subset object contained immature neurons, intermediate progenitor cells, and three classes of neural progenitor cells.

### Fetal brain and COs transcriptome analyses

Single-cell transcriptomic analyses of fetal brain cortex and COs were carried out on a large, publicly available dataset^19^ (downloaded from https://cells.ucsc.edu/organoidreportcard/). The data were handled in Seurat^64^ (v3.2.2) with R version 4.0.2 with a publicly available dataset. The fetal dataset was comprised of 189,409 cells from various cortical regions of five unique fetal donors between weeks 6-22 of gestation. The organoid dataset was comprised of 235,121 cells from 37 organoids across a developmental time window spanning 3-10 weeks of culture. Cell identities were assigned based on the meta data provided by Bhaduri et al.

### Statistical analyses

GraphPad Prism v9.0.0 (La Jolla, CA) was used to generate all graphs and perform all data and statistical analyses in this study. For organoid growth analyses, the number of replicates displayed in Figure 2 ranged from 19-53 from each group. Statistical analysis of data of organoid growth was performed using a Mann-Whitney U test. Statistically significant differences between parental control and proband COs were observed between groups commencing at day 20, p values are displayed in Fig.2d. *p ≤ 0.05; **p ≤ 0.01; ***p ≤ 0.001; ****p ≤ 0.0001; ns=not significant. For organoid growth analyses in Fig.4r, the number of replicates ranged from 44-65 from each group at day 30. Statistical analyses were performed using one-way ANOVA with Tukey’s multiple comparisons test (n = 14 rosettes quantified across a minimum of 4 independent organoids per group and 1 independent experiment performed). Statistical analyses for quantification of EdU+ and pHH3+ NPCs in neural rosettes of organoids was performed using one-way ANOVA with Tukey’s multiple comparisons test (minimum of n = 4 independent organoids per group and 3 independent experiments performed). Cilia length quantifications in neural rosettes of organoids were performed using one-way ANOVA with Tukey’s multiple comparisons test (minimum of n = 4 independent organoids per group and 1 independent experiment performed). Analysis of single-cell RNA sequencing data are described in detail within the methods section. Eight individual organoids were pooled per group consisting of parental control, proband and GC control organoid groups. In all differential expression data analysis and Metascape gene ontology analyses, p values were adjusted for multiple test correction. Significance of differentially expressed genes was defined as adjusted p < 0.05.

## Data Availability

All data produced in the present study are available upon reasonable request to the authors

## Acknowledgements

We are grateful to all laboratory members for their advice and constructive critiques related to this study. We would especially like to thank Arelis Berrios Hester for her excellent editorial assistance. Figures 2 and 4 contain schematics that were created with BioRender.com.

## Author Contributions

S.R.F., W.S., D.J., T.B., and M.E.H. conceived and designed the experiments. S.R.F., W.S., M.E.H. D.J., J.W., T.B., K.E.M., P.W., D.C.K., R.N., and J.F performed the research. S.R.F., M.E.H, W.S., D.J., J.W., T.B., K.E.M., P.W., D.C.K., R.N., S.R., M.N.K., S.E.H., and J.F analyzed the data. S.R.F., W.S., D.J., D.C.K., and M.E.H. wrote the manuscript. All authors provided a critical review and approval of the final manuscript for publication. This bench science and clinical research team works in the Steve and Cindy Rasmussen Institute for Genomic Medicine at Nationwide Children’s Hospital. The institute is generously supported by the Nationwide Foundation Pediatric Innovation Fund.

## Competing interests

The authors report no competing interests.

## Notes

### Competing Interest Statement

The authors have declared no competing interest.

### Funding Statement

The funding of this work was generously supported by the Nationwide Foundation Pediatric Innovation Fund.

### Author Declarations

The IRB committee of Nationwide Children's Hospital gave ethical approval for this work

