## Supplementary Information for "Cerebral Organoids Containing an *AUTS2* Missense Variant Model Microcephaly"

### Supplementary Figure 1

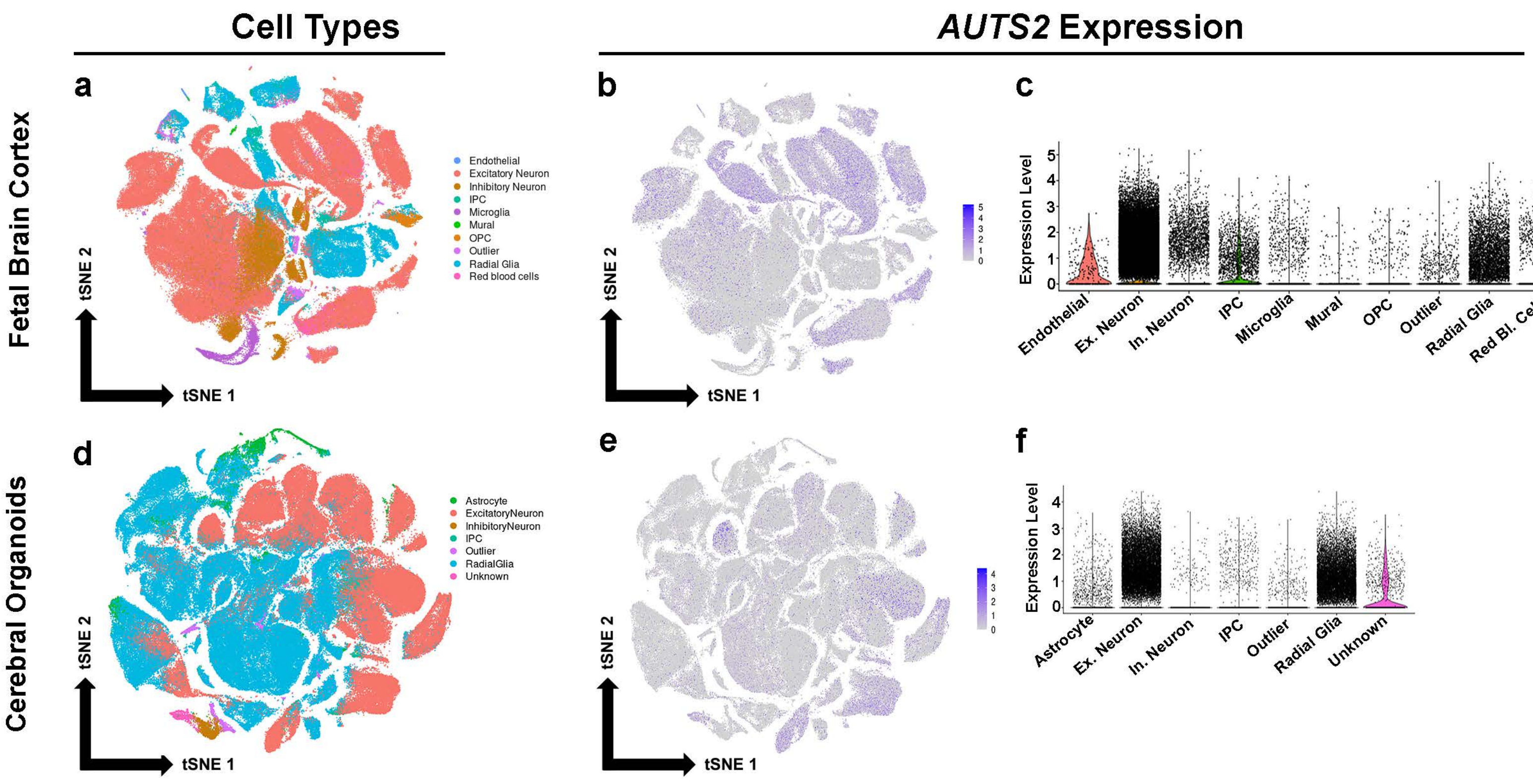

### Supplementary Figure 2

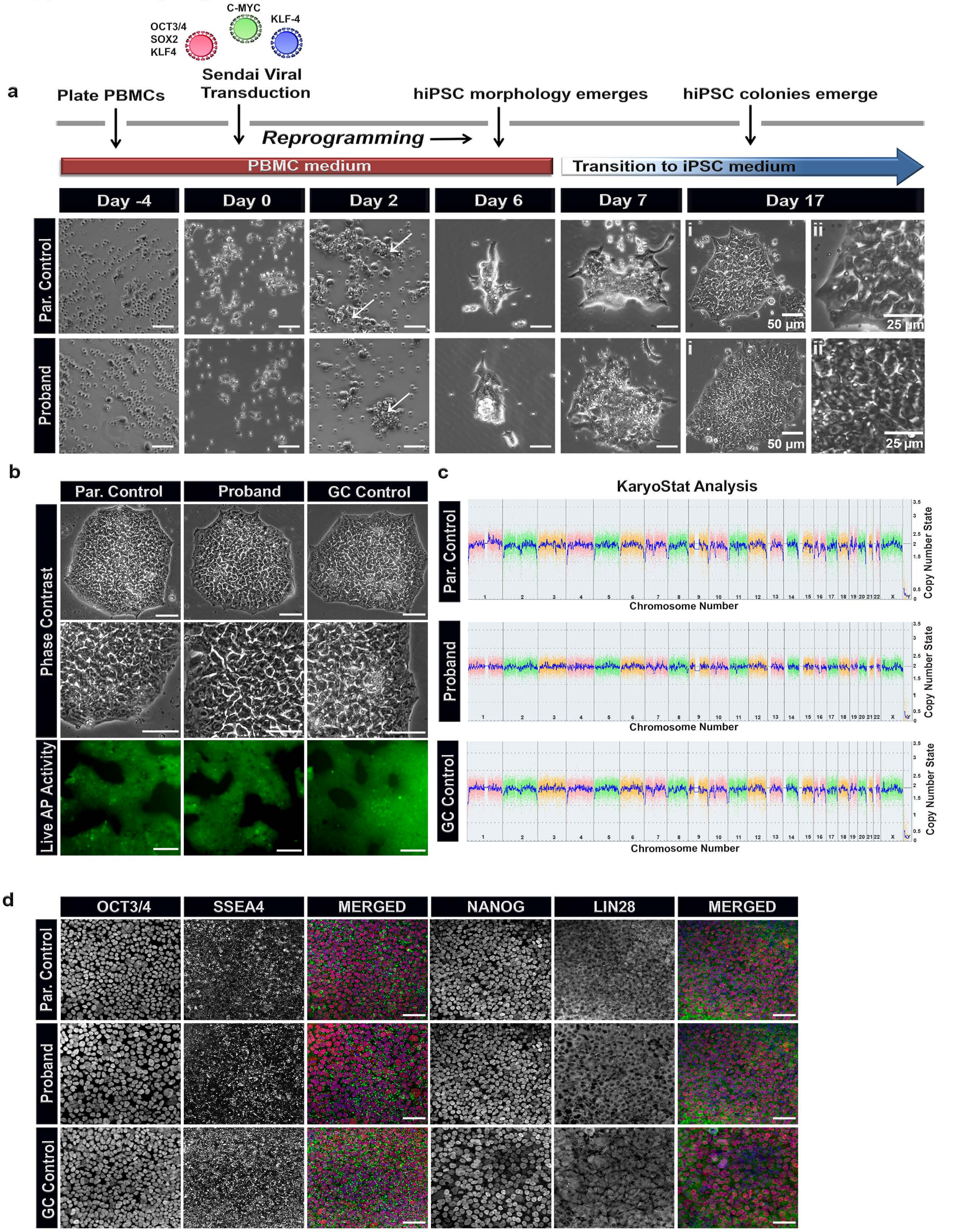

### Supplementary Figure 3

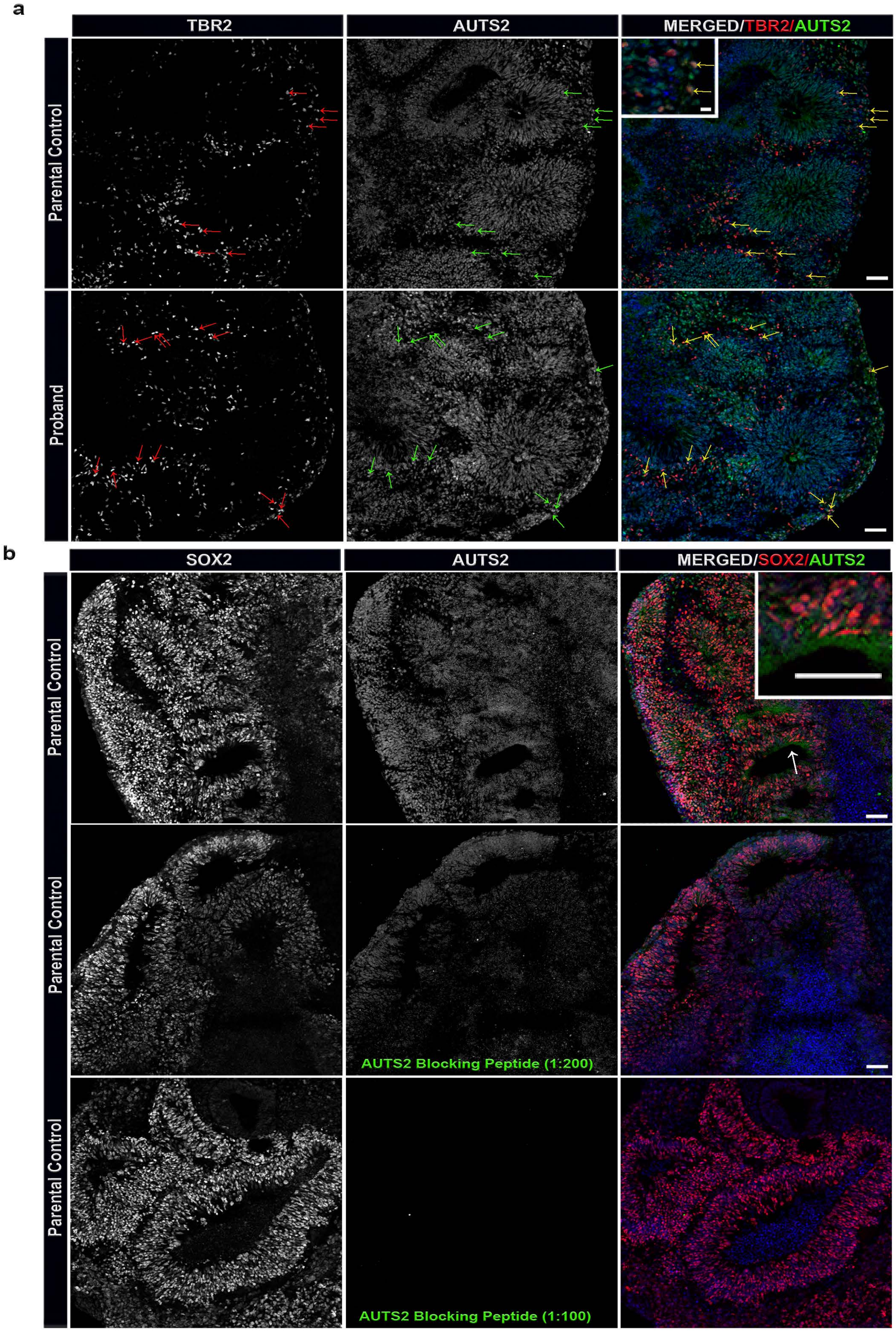

#### Supplementary Figure 4

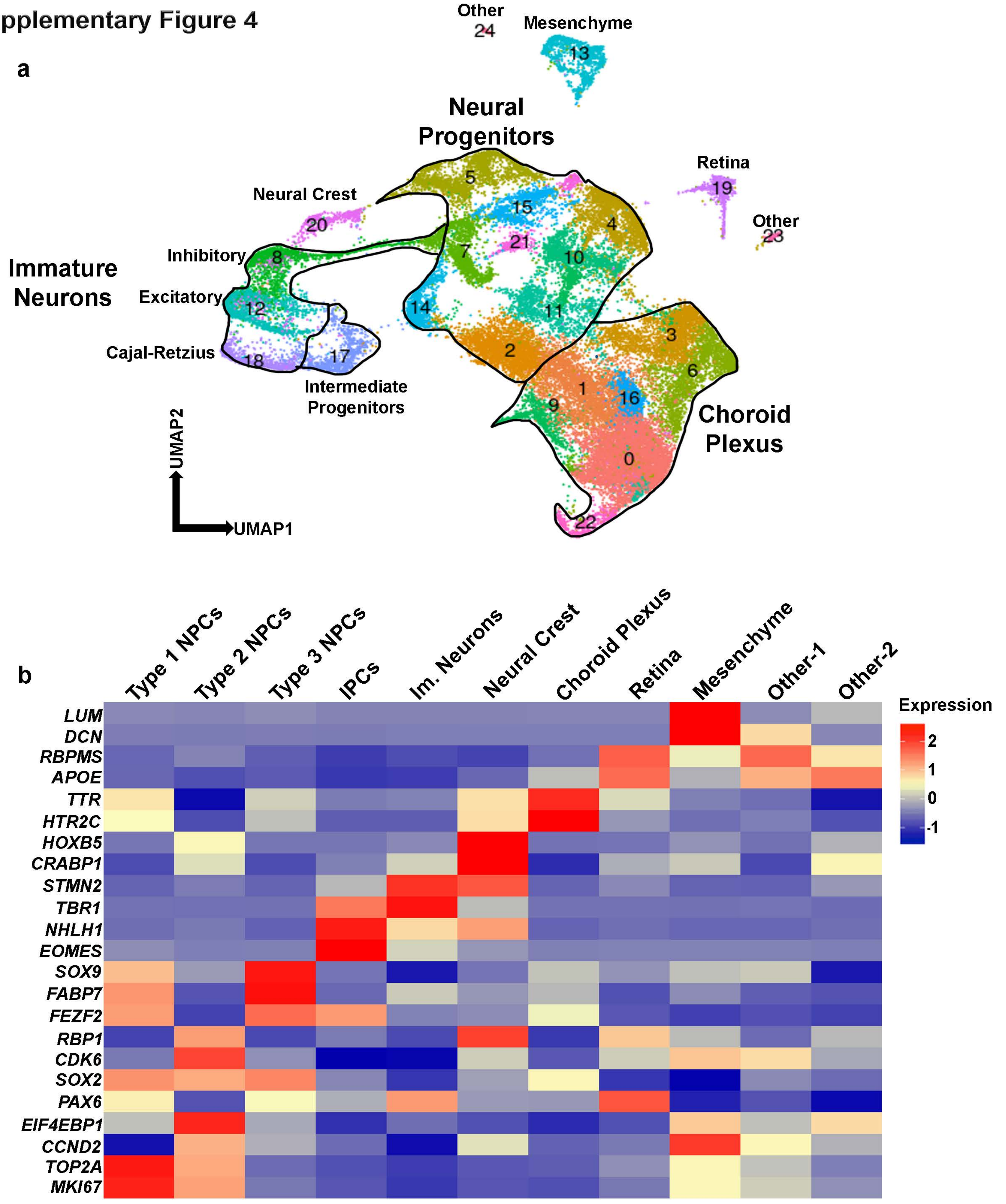

#### Supplementary Figure 5

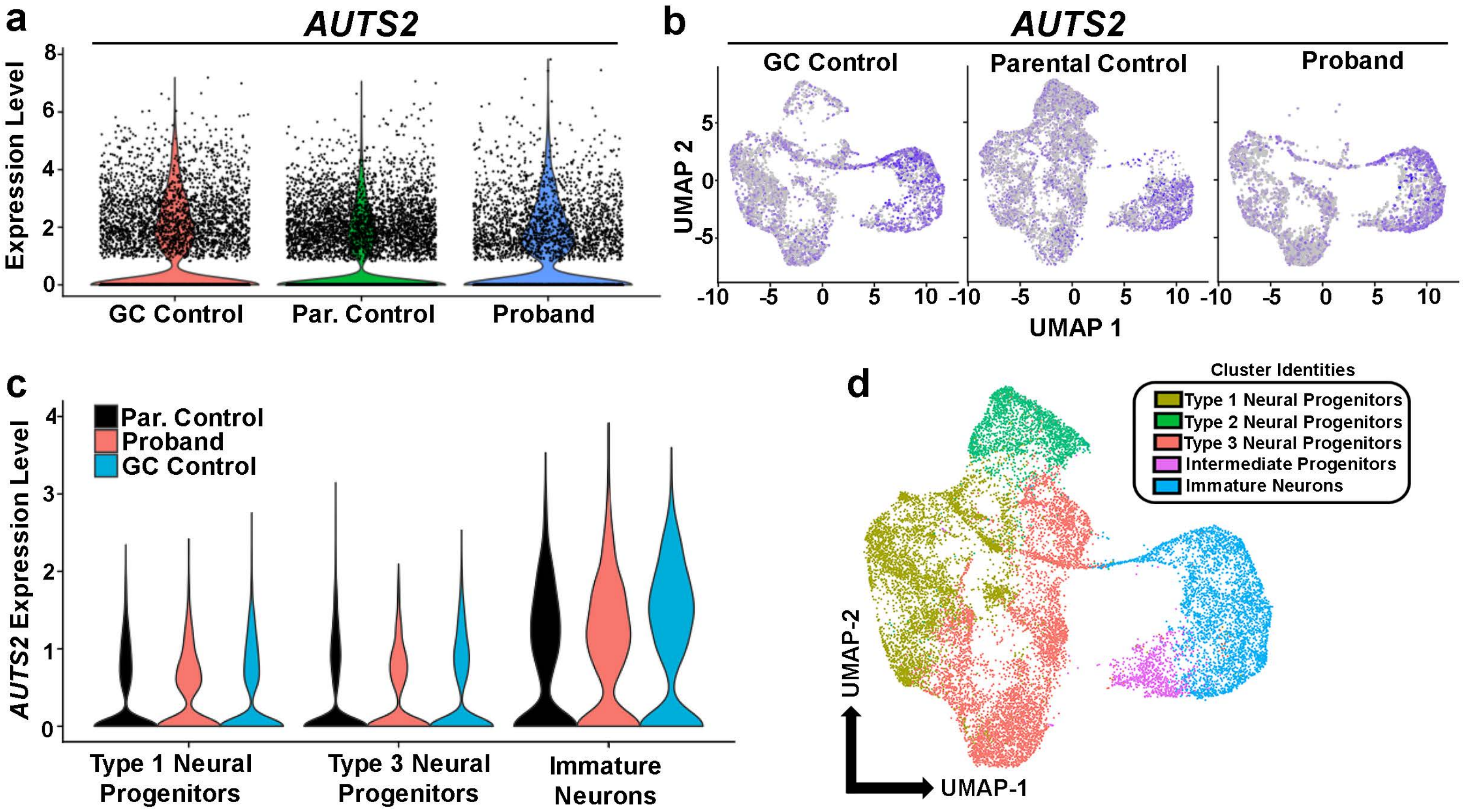

#### Supplementary Figure 6

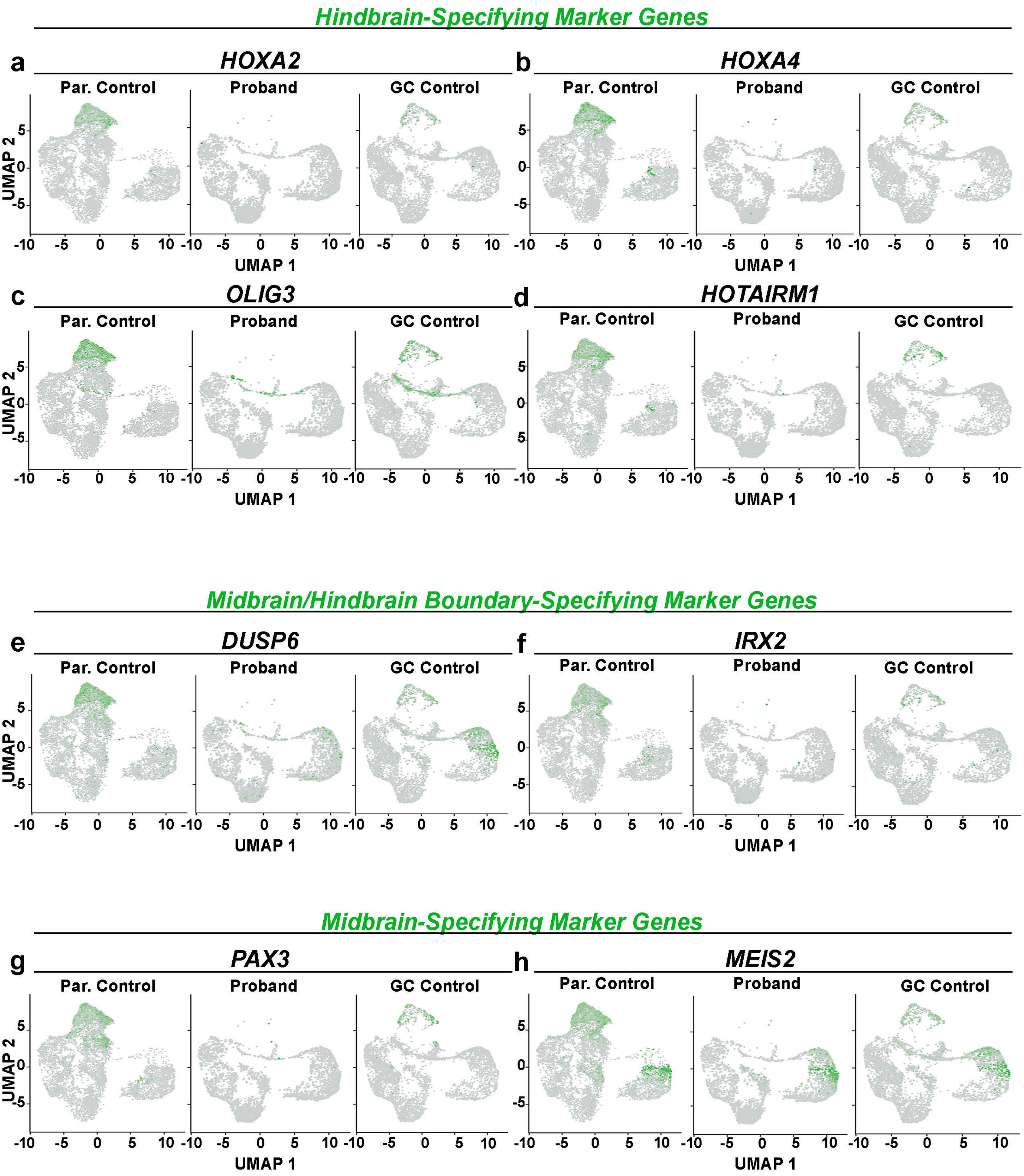

**Supplementary Table 1.** Whole-genome sequencing metrics for the family trio.

| Sample | Read Length | Total Seq. (Gbp) | Map Rate | Dup. Rate | Mean Depth | Cov>=20x |
|---|---|---|---|---|---|---|
| Proband | 150 | 138.3 | 97.6% | 7.6% | 39.07x | 96.50% |
| Parent 1 | 150 | 141.0 | 96.9% | 8.9% | 39.62x | 96.80% |
| Parent 2 | 150 | 279.3 | 49.3% | 10.1% | 39.26x | 95.30% |

**Supplementary Table 2.** Primary antibodies used for immunofluorescence studies.

| Antibody | Full Name | Description | Vendor | Catalog # | Species | Dilution |
|---|---|---|---|---|---|---|
| **NANOG** | Homeobox Transcription Factor Nanog | Pluripotency marker | R&D Sys. | AF1997 | Gt | 1:200 |
| **TPX2** | TPX2 Microtubule Nucleation Factor | Mitotic spindle assembly factor | Novus | NB500-179 | Rb | 1:200 |
| **pHH3** | Phospho-Histone H3 | M-phase specific proliferation marker | Millipore | 06-570 | Rb | 1:500 |
| **ARL13B** | ADP Ribosylation Factor Like GTPase 13B | Cilia marker | Proteintech | 17711-1-AP | Rb | 1:200 |
| **ZO1** | Tight Junction Protein 1 | Apical zone marker | BD Biosciences | 610966 | Ms | 1:300 |
| **SOX2** | SRY-Box 2 | Neural progenitor cell marker | Santa Cruz | sc-17320 | Gt | 1:200 |
| **SSEA4** | Stage-Specific Embryonic Antigen-4 | Pluripotency marker | Millipore | MAB4304 | Ms | 1:200 |
| **LIN28** | LIN-28 family RNA-binding protein | Pluripotency marker | ThermoFisher | PA1-096 | Rb | 1:100 |
| **OCT3/4** | Octamer-Binding Protein 4 | Pluripotency marker | Santa Cruz | SC-8628 | Gt | 1:200 |
| **AUTS2** | Autism Susceptibility Candidate 2/ Activator of Transcription and Developmental Regulator | Transcriptional regulator protein | Sigma | HPA000390 | Rb | 1:200 |
| **AUTS2 blocking peptide** | PrEST Antigen AUTS2 | AUTS2 blocking peptide (30kDA) | Santa Cruz | APREST76 148 | N/A | 1:100, 1:200 |
| **TBR2** | T-Box, Brain 2 | Intermediate progenitor (IP) marker | Abcam | Ab23345 | Rb | 1:300 |

**Supplementary Table 3.** sgRNA and ssODN sequences used for CRISPR/Cas9 genome editing.

| | |
|---|---|
| **Modification** | P534T (CCC>ACC) |
| **Guide RNA Sequence** | TGTGCTGGTGCGTGTGCTGG |
| **Guide RNA cut location** | chr7:70,766,253 |
| **Donor Sequence** | CGCCCTACCTGCGGACCGAGTTCCATCAGCACCAGC<br>ACCAGCACCAGCACACCCACCAACACACGCACCAG<br>CACACCTTCACGCCGTTCCCCCACGCCATCCC |

**Supplementary Table 4.** Cerebral Organoid Media Formulations.

| Neural Induction Media | | |
|---|---|---|
| **Product** | **Vendor/ Catalog No.** | **Amount** |
| DMEM-F12 | ThermoFisher #11320033 | 250.0 **mL** |
| MEM-NEAA | ThermoFisher #11140-050 | 2.5 **mL** |
| GlutaMAX Supplement | ThermoFisher #35050061 | 2.5 **mL** |
| N-2 Supplement | ThermoFisher #17502-048 | 2.5 **mL** |
| Anti-Anti | ThermoFisher #15240062 | 2.5 **mL** |

| Cerebral Organoid Expansion Medium | | |
|---|---|---|
| **Product** | **Vendor/ Catalog No.** | **Amount** |
| StemCell BrainPhys™ | Stem Cell Technologies #05792 | 250 **mL** |
| MEM-NEAA | ThermoFisher #11140-050 | 1.25 **mL** |
| SM1 without Vitamin A | Stem Cell Technologies #05731 | 2.5 **mL** |
| N-2 Supplement | ThermoFisher #17502-048 | 1.25 **mL** |
| GlutaMAX Supplement | ThermoFisher #35050061 | 2.5 **mL** |
| Anti-Anti | ThermoFisher #15240062 | 2.5 **mL** |
| Insulin | Sigma #I9278 | 62.5 **µL** |
| Matrigel | Corning #354230 | 5.0 **mL** |

| Cerebral Organoid Growth & Differentiation Media | | |
|---|---|---|
| **Product** | **Vendor/ Catalog No.** | **Amount** |
| StemCell BrainPhys™ | Stem Cell Technologies #05790 | 250 **mL** |
| MEM-NEAA | ThermoFisher #11140-050 | 1.25 **mL** |
| GlutaMAX Supplement | ThermoFisher #35050061 | 2.5 **mL** |
| SM1 with Vitamin A | Stem Cell Technologies #05711 | 2.5 **mL** |
| N-2 Supplement | ThermoFisher #17502-048 | 1.25 **mL** |
| Anti-Anti | ThermoFisher #15240062 | 2.5 **mL** |
| Insulin | Sigma #I9278 | 62.5 **µL** |
| **Dilute (1:100)** 2-BME Solution | Sigma #M-3148 | 87.5 **µL** |
| Matrigel | Corning #354230 | 2.5 **mL** |
